# Observational and genetic associations between cardiorespiratory fitness and cancer: a UK Biobank and international consortia study

**DOI:** 10.1101/2023.03.28.23287805

**Authors:** Eleanor L. Watts, Tomas I. Gonzales, Tessa Strain, Pedro F. Saint-Maurice, D. Timothy Bishop, Stephen J. Chanock, Mattias Johansson, Temitope O. Keku, Loic Le Marchand, Victor Moreno, Polly A. Newcomb, Christina C. Newton, Rish K. Pai, The PRACTICAL consortium, CRUK, BPC3, CAPS, PEGASUS, Mark P. Purdue, Cornelia M. Ulrich, Karl Smith-Byrne, Bethany Van Guelpen, Felix R. Day, Katrien Wijndaele, Nicholas J. Wareham, Charles E. Matthews, Steven C. Moore, Soren Brage

**Author notes:** Corresponding author Eleanor L. Watts, D.Phil. Metabolic Epidemiology Branch, Division of Cancer Epidemiology and Genetics, National Cancer Institute, National Institutes of Health, Rockville, Maryland, 20850, USA. ORCID: 0000-0001-9229-2589. These authors contributed equally to this work (Eleanor L. Watts, Tomas I. Gonzales). Joint senior authors (Steven C. Moore, Soren Brage). Members from the PRACTICAL Consortium, CRUK, BPC3, CAPS and PEGASUS are provided in Appendix 1.

## Abstract

**Importance:** The association of cardiorespiratory fitness with cancer risk is not clear.

**Objective:** To investigate whether fitness is associated with the risk of diagnosis of common cancers.

**Design, setting, and participants:** In observational analyses, we used multivariable-adjusted Cox proportional hazards models to estimate hazard ratios (HRs) and 95% confidence intervals (CIs) for risk of cancer in a subset of UK Biobank participants who completed a submaximal fitness test in 2009-12 (*N*=72,572). In secondary analyses, we used a two-sample Mendelian randomization (MR) framework, with genetically predicted fitness as an instrumental variable derived from UK Biobank study participants and genetic cancer data from international consortia. Odds ratios (ORs) were estimated using the inverse-variance weighted method.

Relationships between fitness and cancer may be partially mediated by adiposity, and therefore associations were estimated with and without adjustment for adiposity.

**Exposures:** Estimated maximal cardiorespiratory fitness (ml O_2_⋅min^-1^⋅kg^-1^ total-body mass and ml O_2_⋅min^-1^⋅kg^-1^ fat-free mass).

**Main outcomes and measures:** Diagnosis of lung, colon, rectal, endometrial, female breast, and prostate cancer. MR analyses additionally included pancreatic and renal cancers.

**Results:** After a median of 11 years of follow-up, 4,290 cancers of interest were diagnosed. A 3.5 ml O_2_⋅min^-1^⋅kg^-1^ total-body mass increase in fitness (approximately 0.5 standard deviation (SD)) was associated with lower risks of endometrial (HR=0.81, 95% CI 0.73-0.89), colorectal (0.94, 0.90-0.99), and breast cancer (0.96, 0.92-0.99). In MR analyses, higher levels of genetically predicted fitness were associated with a lower risk of breast cancer (OR per genetically predicted 0.5 SD increase in ml O_2_⋅min^-1^⋅kg^-1^ fat-free mass=0.92, 95% CI 0.86-0.98), including estrogen receptor (ER)+ (0.91, 0.84-0.99) and ER-(0.88, 0.80-0.97) subtypes. After adjusting for body fat, both the observational and genetic associations were attenuated.

**Conclusions and relevance:** Higher fitness levels may reduce risks of endometrial, colorectal, and breast cancer, though relationships with adiposity are complex and may mediate these relationships. Aiming to increase fitness, including via changes in body composition, may be an effective strategy for cancer prevention.

**KEY POINTS:** **Question:** Is cardiorespiratory fitness associated with subsequent risk of cancer diagnosis?

**Findings:** In a prospective cohort study of 73,000 cancer-free participants who completed a submaximal fitness test, we report that higher fitness levels were associated with lower risks of endometrial, colorectal, and breast cancer. Using two-sample Mendelian randomization methods we also found an inverse association with breast cancer. Associations were attenuated following adjustment for adiposity.

**Meaning:** Higher fitness may be associated with reduced risk of certain cancer sites. Aiming to increase fitness, including via changes in body composition, may be an effective strategy for cancer prevention. The role of adiposity in mediating the relationship between fitness and cancer risk is not fully understood, and further research is needed to explore this complex relationship.

## INTRODUCTION

Until recently epidemiological studies have largely focused on the role of physical activity behaviours with cancer risk^1^. Cardiorespiratory fitness (referred to here as ‘fitness’) is distinct from physical activity as it describes the capacity of the circulatory and respiratory systems to supply oxygen to skeletal muscle during prolonged physical activity^2, 3^. Fitness is generally objectively measured and has a stronger genetic component than habitual physical activity^2–4^.

Higher fitness is associated with good cardiometabolic health, including lower visceral adipose tissue, inflammation and insulin sensitivity, and may therefore reduce the risk of cancer^5–8^. Previous studies report that people with higher fitness have lower risks of all-cause mortality, cancer mortality and cardiovascular disease^5, 9–11^, but the relationship between fitness and incident cancers are less clear. Some studies have reported inverse associations between fitness and lung and colorectal cancers ^12–15^, while for prostate cancer associations have been reported to be null or positive^13–18^. Only one prior study has investigated associations between fitness and female-specific incident cancers, and did not find evidence of a relationship^14^.

A limitation of observational epidemiological studies includes the possibility of residual confounding and reverse causation. Mendelian randomization (MR) uses germline genetic variants as proxies of biological traits to generate instrumental variables and estimate their associations with disease risk. Because germline genetic variants are fixed and randomly allocated at conception, this technique may be less likely to be affected by biases and confounding factors (such as preclinical disease and smoking history). This is the first study to use MR methods to investigate fitness and cancer risk.

We aimed to assess the associations of measured fitness and risk of common cancers (lung, colon, rectal, endometrial, female breast, and prostate cancer) using observational methods in the UK Biobank. In secondary analyses, we used a two-sample MR framework, using genetically predicted fitness, as instrumental variable derived from UK Biobank^19^ and genetic case control data from consortia for those same sites, plus pancreatic cancer and renal cell carcinoma for which observational analyses in the UK Biobank are underpowered. By integrating evidence from both observational epidemiology and MR approaches, we aim to strengthen the basis for causal inference^20^. As excess adiposity may act as a mediator, we examined associations between fitness and cancer with and without adjustment for adiposity.

## METHODS

### UK Biobank study population

The UK Biobank study is a population-based prospective cohort study of 502,625 adults aged 40 to 69 years. A description of the study protocol is available online^21^. Participants were registered with the UK National Health Service and lived within 40 km of a UK Biobank assessment centre in England, Wales, and Scotland. Baseline data were collected between 2006 and 2010. A repeat-measures substudy was conducted between 2012 and 2013. The study was approved by the North West Multicentre Research Ethics Committee. Participants provided written informed consent.

### UK Biobank cardiorespiratory fitness assessment

An individualised submaximal cycle ergometer test was implemented in 2009 and offered to 75,087 participants during baseline data collection, 17,109 participants during the repeat assessment study, and 2,877 participants at both timepoints; 97,950 tests were offered in total.

For those participants who were offered a test at both timepoints, the earliest fitness test completed by the participant was used to maximise follow-up duration. Participant baseline data were collected on the same day as their exercise test. The test was individualized to each participant’s exercise capacity and risk level for engaging in exercise. Participants with lower exercise capacity or higher risk for exercise-related complications were offered a test with lower work rates, while those with higher exercise capacity or lower risk were offered a test with higher work rates. A description of the exercise test individualisation process and maximal oxygen consumption (VO_2_ max; ml O_2_⋅min^-1^⋅kg^-1^) estimation process is provided in supplemental methods; the test protocol is available online.^22^ VO_2_ max was estimated in two ways: scaled by total-body mass (VO2maxtbm [3.5 ml O2⋅min^-1^⋅kg^-1^ total-body mass=1 MET]) and scaled by fat-free mass (VO2maxffm).^23, 24^ VO2maxffm represents the ability of skeletal muscle to use oxygen during maximal exercise, whereas VO2maxtbm is more representative of aerobic performance capacity.^25^

### Genetic instrument for cardiorespiratory fitness

Full details of the fitness genome-wide association study (GWAS) are available elsewhere.^19^ In brief, single nucleotide polymorphisms (SNPs) associated with fitness were identified from a GWAS based on UK Biobank participants of European ancestry who participated in the fitness test (*N* included=69,416). Fitness was estimated using the same framework method described above, scaled by fat-free mass and using resting heart rate data from the full cohort, excluding those taking beta-blockers (*N* included=452,941) (*P*<5 x 10^-8^ significance threshold).

The Radial plot method was used to select eligible resting heart-rate associated genetic variants for fitness by removing heterogeneous outliers for the genetic variants, of which 149 were also nominally significant in the fitness GWAS (p<0.05)^26^. The genetic instrument for fitness included 14 fitness and 149 fitness and resting heart rate variants with prioritisation given to the variants identified in the fitness GWAS. In total, 160 independent (r²> 0.01) genetic variants were included in our instrument for fitness.^19^

### Cancer ascertainment

#### Observational analysis

Both cancer registration data and Hospital Episode Statistics (HES) were used to identify participants with incident cancer (see Supplemental Methods for cancer site definitions). Cancer registration data were provided via record linkage to national cancer and death registries, until the following censoring dates: 31 July 2019 in England and Wales and 31 October 2015 in Scotland. Cancers occurring after the registry censoring dates were identified using HES data, until the following censoring dates: 30 September 2021 in England, 31 July 2021 in Scotland and 28 February 2018 in Wales.

Of the 84,792 fitness tests analysed after preliminary exclusions (i.e. participant withdrawal of data, “high risk” for exercise; see Supplementary Figure 1), we retained a preliminary analytic sample of 79,347 participants after additionally excluding 3,209 participants for missing data, 1,017 due to test data quality, 1,219 with missing weight, fat-free mass, or heart rate, and 44 for whom fitness estimation could not be applied. We then excluded 5,180 participants with prevalent cancer at baseline and 1,551 participants diagnosed with cancer within two years of follow-up. The final analytic sample was 72,572 participants. Health and sociodemographic characteristics were described across age-adjusted and sex-specific fitness tertiles^27^.

#### Genetic cancer data

Risk estimates may be biased when instrumental variables and outcomes are identified from the same sample^28^. We therefore used independent GWAS data from international consortia. This includes breast (including estrogen receptor (ER)+ and ER-subtypes)^29, 30^, prostate (including aggressive disease)^31^, endometrial^32^, ovarian^33^, lung (including for never smokers)^34^, and colorectal cancer (including colon, rectal, male colorectal and female colorectal, distal colon and proximal colon)^35, 36^. We also included pancreatic cancer and renal cell carcinoma^37–40^. Included sites and subtypes were chosen based on data availability. Further information for the genetic case control studies for each cancer site are available from Supplementary Table 1.

### Statistical analysis

#### Observational analysis

Cox regression models with age as the underlying timescale were used to estimate hazard ratios (HRs) and 95% confidence intervals (CIs) per 3.5 ml⋅O_2_⋅min^−1^⋅kg^−1^ total-body mass and 5.0 ml⋅O_2_⋅min^−1^⋅kg^−1^ fat-free mass for risk of cancer diagnosis. Models were progressively adjusted for possible confounding factors (see Supplemental Methods). Adiposity may partially mediate and confound the relationship between fitness and cancer risk (Supplementary Figure 2). Therefore, we evaluated the role of adiposity in fitness-to-cancer associations both with and without adjustment for either BMI (for models with VO_2_max scaled by total-body mass) or fat mass (for models with VO_2_max scaled by fat-free mass).

We have shown previously that repeat assessments of the UK Biobank fitness test will elicit moderately stable fitness estimates (regression dilution ratio=0.79, standard error=0.01)^42^. This source of measurement error will influence the strength of observed health associations.

Therefore, in a sensitivity analysis, we also provide regression dilution calibrated estimates of fitness-to-cancer associations using established statistical techniques^43^. The shape of dose-response relationships between fitness and risk of cancer diagnosis was investigated using cubic spline regression models. Each model used two knots placed at the 33^rd^ and 67^th^ percentile of the fitness distribution. Reference values were set to the mean fitness value for each specific analysis (see Supplementary Figures 3 and 4).

##### Sensitivity analysis

Subgroup analyses for colorectal cancer were examined by sex, and associations for fitness and lung cancer were re-examined after restricting the analysis to never-smokers only. Subgroups were chosen *a priori* on the basis of data availability and previous evidence for heterogeneity in the associations^1^. We also included a minimally adjusted model to investigate the influence of mediators and/or confounders.

#### Mendelian randomization

The MR estimation for fitness and cancer was conducted using the inverse-variance weighted (IVW) method^44^. We additionally calculated the I^2^_GX_ statistic to assess measurement error in SNP-exposure associations^31^, the F-statistic to examine the strength of the genetic instrument^45^, Cochran’s Q statistic for heterogeneity between the MR estimates for each SNP ^46^, and PhenoScanner was used to assess pleiotropy of the genetic instruments^47^. As sensitivity analyses, we used the MR residual sum and outlier (MR-PRESSO) and leave-one out analyses to investigate the role of SNP outliers^48^. To assess pleiotropy, we used the weighted median and contamination mixture methods^49^.

To explore relationships between body fat and fitness, we conducted a bi-directional MR of genetically predicted fitness on fat mass and *vice versa* using our genetic instrument for fitness and an instrument for total fat mass based on a GWAS of UK Biobank participants (*N*=330,762 participants of European ancestry), derived from bioelectrical impedance measurements at study baseline^50^. We also conducted multivariable MR (MVMR) analyses to assess the effect of fitness on cancer risk, after accounting for genetically predicted fat mass and height^44^.

#### Statistical software

Observational analyses were performed using Stata version 16.1 (Stata Corporation, College Station, TX, USA). MR analyses were performed using the *TwoSampleMR* and *MendelianRandomisation* R packages^51, 52^ and figures were plotted in R version 3.6.3. All tests of significance were two-sided, and P-values <0.05 were considered statistically significant. Results are presented in accordance with the STROBE checklist.

## RESULTS

### Observational analysis

After a median of 11 years of follow-up, 1,586 prostate cancers, 1,093 breast cancers, 811 colorectal cancers, 480 lung cancers, 184 endometrial cancers, and 136 ovarian cancers were diagnosed. Participant characteristics by age-adjusted and sex-specific fitness tertiles are provided in **Table 1** for fitness scaled by total-body mass and Supplementary Table 2 for fitness scaled by fat-free mass. Fitness was higher in men compared to women, and those in the middle and higher fitness tertiles had better measures of adiposity, socioeconomic status, and cardiometabolic health than those in the lower fitness tertile.

**Table 1:**
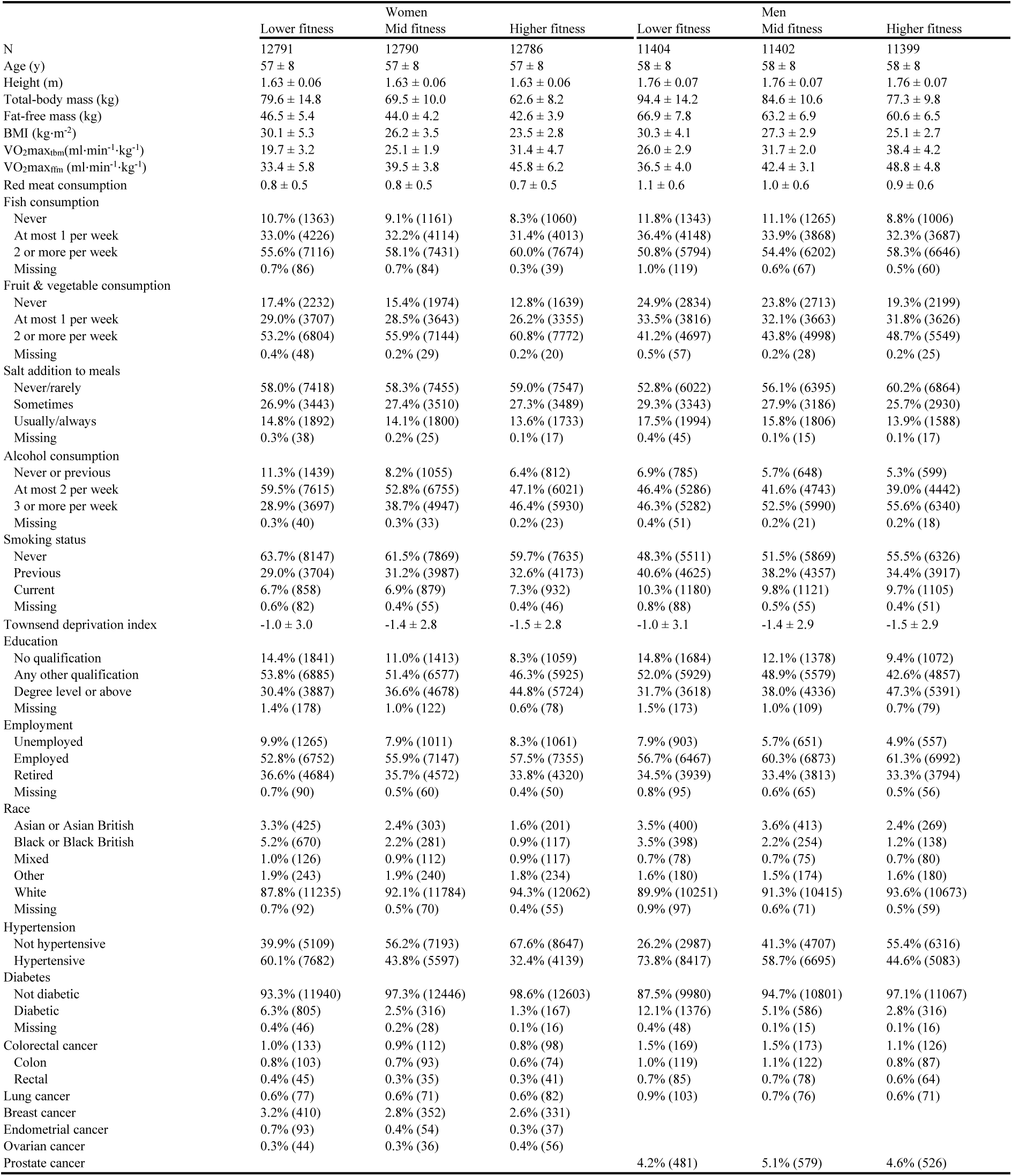
Participant characteristics by age-adjusted and sex-specific cardiorespiratory fitness (VO2max per kg total-body mass) tertiles

Observational analysis results are summarised in **Figure 1**. In analyses without BMI adjustment, each 3.5 ml O_2_⋅min^-1^⋅kg^-1^ total-body mass increase (equivalent to 1 metabolic equivalent of task [MET]) in fitness was associated with a 19% reduction in endometrial cancer, 6% reduction in colorectal cancer, and 4% reduction in breast cancer. After BMI adjustment, associations were attenuated but remained directionally consistent. Where associations were detected, relationships generally appeared to be linear but with uncertainty for some cancers at the tails of the fitness distribution (Supplementary Figures 3 and 4). When fitness was expressed per kg fat-free mass, associations with cancers were not significant. Results adjusted for regression dilution are shown in Supplementary Figure 5; unadjusted and adjusted results were generally similar, and the statistical significance of associations were unchanged.

**Figure 1:**
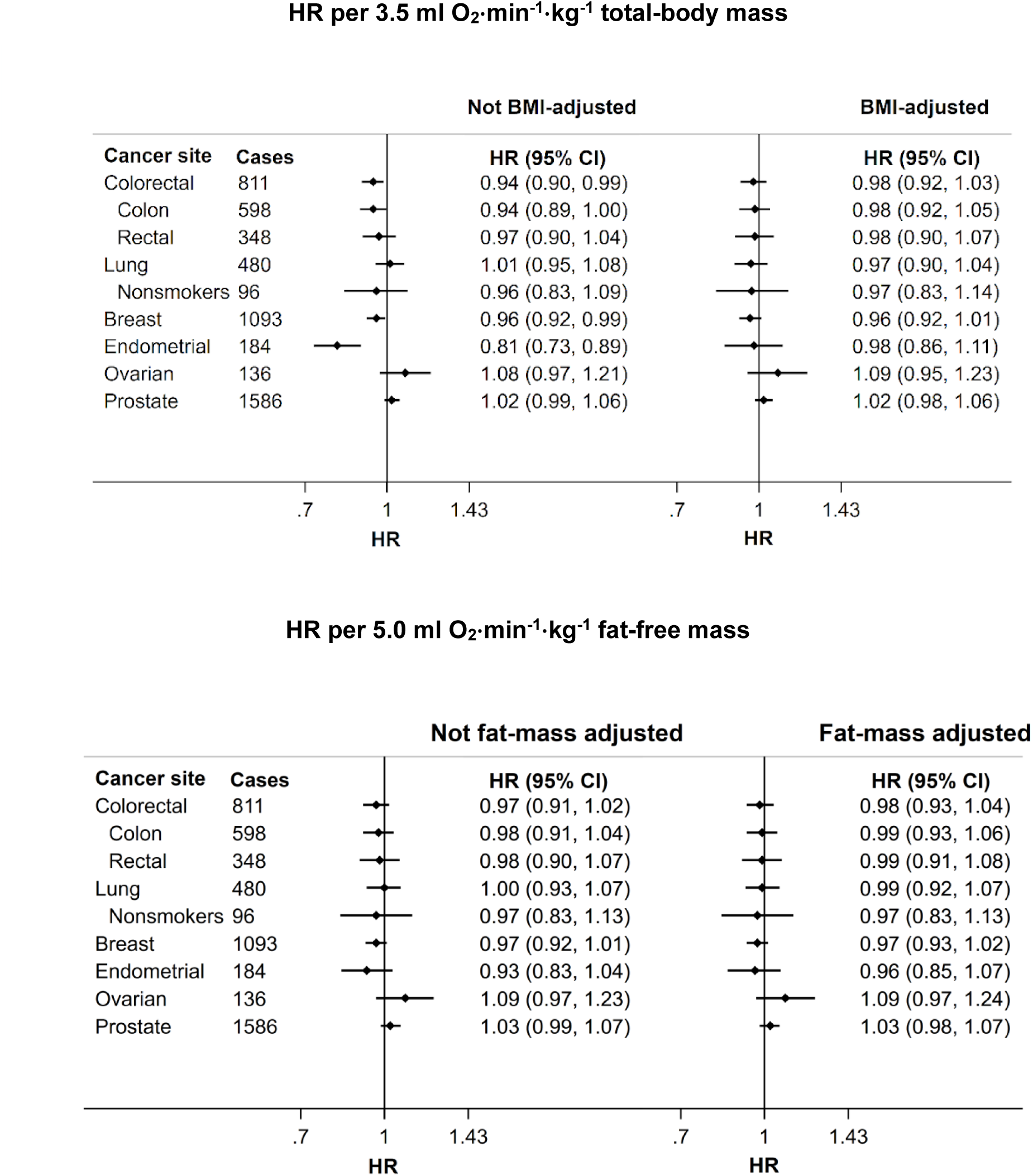
Associations of cardiorespiratory respiratory fitness and incident cancer risk without and with body fat adjustment HRs and 95% CIs estimated using Cox regression models adjusted for age, sex, self-reported racial/ethnic group, Townsend index of deprivation, education, employment status, smoking status, alcohol consumption, red and processed meat consumption, fish consumption, fruit and vegetable consumption, salt consumption, diabetes status, hypertension, medication use (beta blockers, calcium channel blockers, ACE inhibitors, diuretics, bronchodilators, lipid-lowering agents, iron deficiency agents, non-steroidal anti-inflammatory drugs, metformin). Female reproductive cancers (breast, endometrial, and ovarian) were additionally adjusted for age at menarche, age at menopause, parity, hormone replacement therapy usage, and oral contraceptives. Associations with and without adjustment for either continuous BMI (for models with VO_2_max scaled by total-body mass) or fat mass (for models with VO_2_max scaled by fat-free mass). Abbreviations: ACE=Angiotensin-converting enzyme; BMI=body mass index; CI=confidence interval; HR=hazard ratio.

There was evidence of heterogeneity in the associations of fitness and colorectal cancers by sex; the relationship was inverse for men and null for women (**Figure 2** and Supplementary Figure 4). Minimally adjusted models are available from Supplementary Table 3.

**Figure 2:**
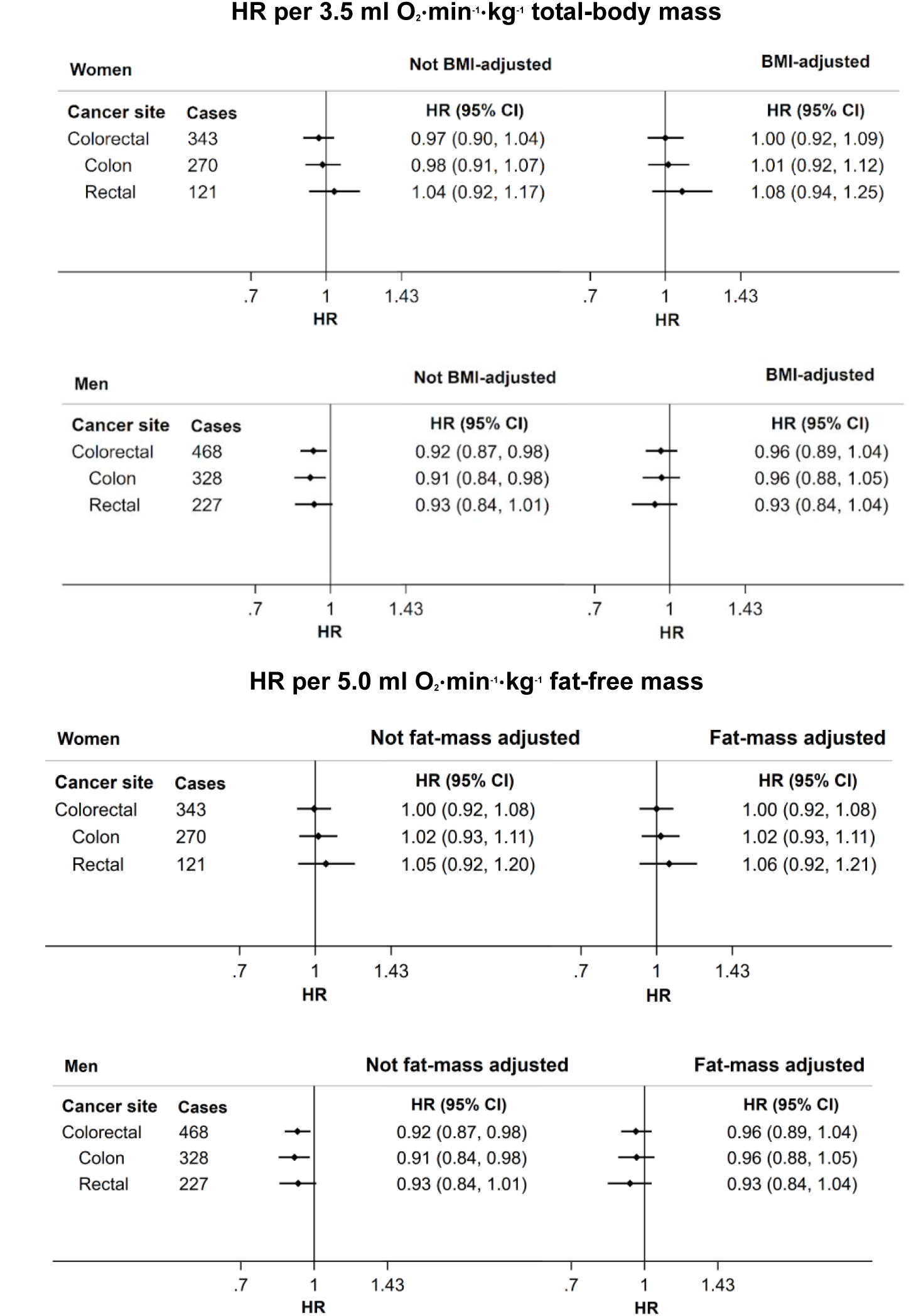
Sex-stratified associations of cardiorespiratory respiratory fitness and incident cancer risk without and with body fat adjustment HRs and 95% CIs estimated using Cox regression models adjusted for age, sex, self-reported racial/ethnic group, Townsend index of deprivation, education, employment status, smoking status, alcohol consumption, red and processed meat consumption, fish consumption, fruit and vegetable consumption, salt consumption, diabetes status, hypertension, medication use (beta blockers, calcium channel blockers, ACE inhibitors, diuretics, bronchodilators, lipid-lowering agents, iron deficiency agents, non-steroidal anti-inflammatory drugs, metformin). Associations with and without adjustment for either continuous BMI (for models with VO_2_max scaled by total-body mass) or fat mass (for models with VO_2_max scaled by fat-free mass). Abbreviations: ACE=Angiotensin-converting enzyme; BMI=body mass index; CI=confidence interval; HR=hazard ratio.

### Mendelian randomization analyses

Higher levels of genetically predicted fitness were associated with a lower risk of breast cancer (OR per 5.0 ml O_2_⋅min^-1^⋅kg^-1^ fat-free mass=0.92, 95% CI 0.86-0.98; P=0.02), including ER+ (0.91, 0.84-0.99; P=0.02) and ER- (0.88, 0.80-0.97; P=0.01) subtypes, but was not significantly associated with any other cancer site (**Figure 3**). There was also no evidence of an association with colorectal cancer after stratification by sex and site (Supplementary Tables 4 and 5). There was significant heterogeneity in the MR estimates for the SNPs for each cancer site (Cochran’s Q P<0.05), except for associations with lung cancer for never smokers (P=0.13), aggressive prostate cancer (P=0.17) and renal cancer (P=0.09).

**Figure 3:**
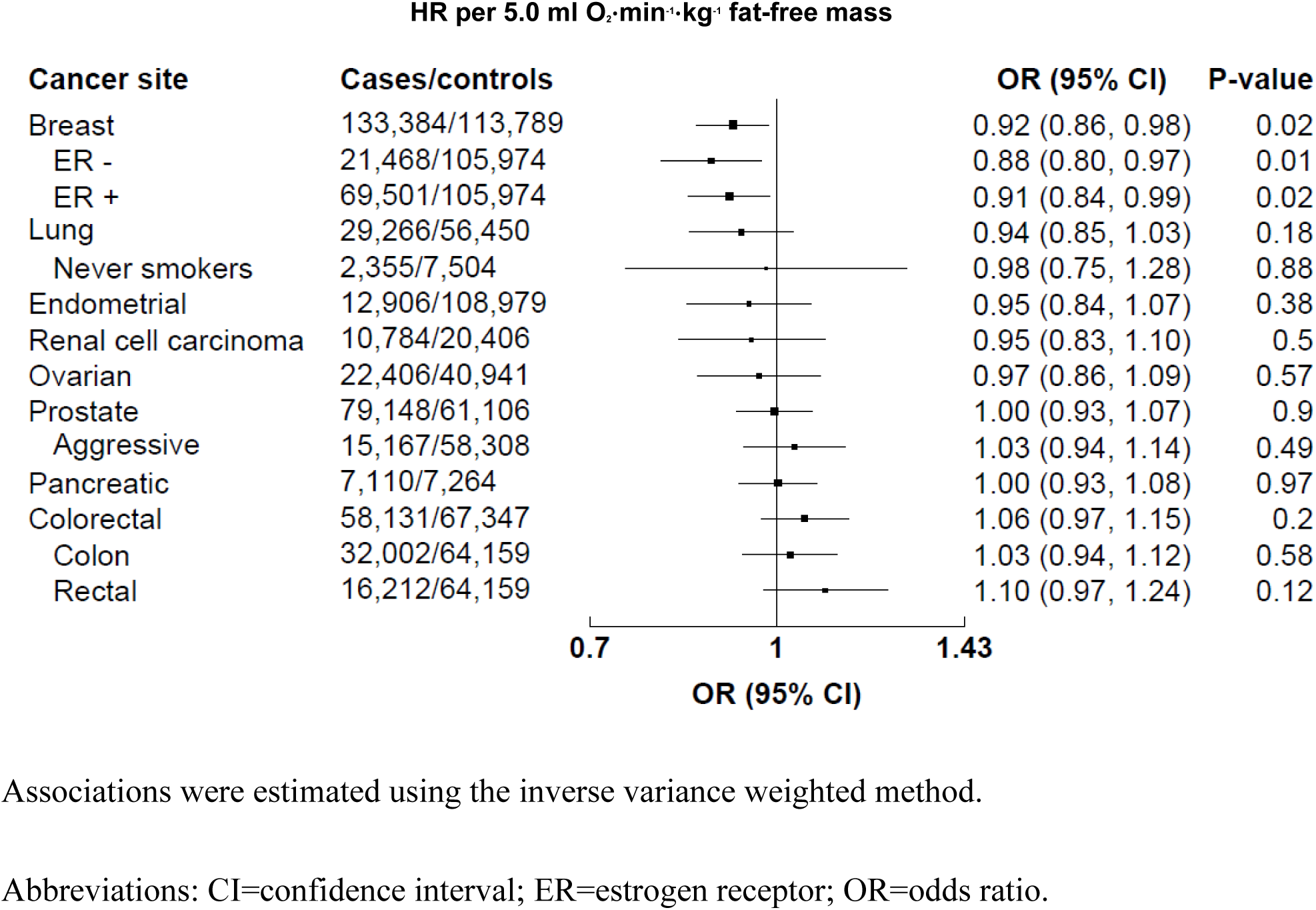
Associations of genetically predicted cardiorespiratory respiratory fitness and cancer risk Associations were estimated using the inverse variance weighted method. Abbreviations: CI=confidence interval; ER=estrogen receptor; OR=odds ratio.

In MR sensitivity analyses, the relationships between fitness and breast cancer were directionally consistent in comparison with the primary MR analysis (Supplementary Table 6). There was evidence of an inverse association between fitness and lung cancer using the weighted median method (0.85, 0.74-0.98; P=0.02) and a positive association with pancreatic cancer using the contamination mixture method (1.09, 1.03-1.14; P=0.03), but no other relationship was statistically significance (Supplementary Table 6). Radial plots also did not indicate any strong influence of outliers on the MR results (Supplementary Figure 6). The likelihood of bias due to weak instruments was low (F-statistic > 10 for all SNPs). There was evidence of moderate levels of measurement error (I^2^_GX_=0.52-0.65), indicating reduced reliability of Egger results, therefore we do not include Egger estimates^53^. Using PhenoScanner, 742 traits were linked to SNPs for fitness (P<5 x 10^-8^), particularly pulse rate (Supplementary Figure 7).

The bi-directional MR analysis indicated that genetically instrumented fat mass had a strong inverse association with fitness (OR per 0.5 SD increase=0.61, 0.52-0.71; P<0.001), but a weaker inverse relationship of fitness with fat mass (OR per 5 ml O_2_⋅min^-1^⋅kg^-1^ fat-free mass=0.96, 0.92-1.01; P=0.08). In MVMR analyses, associations with breast cancer were attenuated after adjustment for fat-free mass and height. While associations with lung cancer became statistically significant (0.90, 0.84-0.96; P=0.002), but genetic associations were null for never smokers **(Table 2**).

**Table 2:** Genetic associations of cardiorespiratory respiratory fitness and cancer risk after accounting for fat mass and height

| Cancer site | OR per 5.0 ml O<sub>2</sub>·min<sup>-1</sup>·kg<sup>-1</sup><br>fat-free mass (95% CI) | P-value |
| --- | --- | --- |
| Breast | 0.98 (0.93, 1.03) | 0.39 |
| ER - | 0.95 (0.89, 1.02) | 0.14 |
| ER + | 0.97 (0.92, 1.03) | 0.36 |
| Lung | **0.90 (0.84, 0.96)** | **0.002** |
| Never smokers | 0.98 (0.82, 1.17) | 0.83 |
| Endometrial | 0.97 (0.89, 1.06) | 0.24 |
| Renal cell carcinoma | 0.86 (0.73, 1.01) | 0.07 |
| Ovarian | 1.00 (0.92, 1.08) | 0.99 |
| Prostate | 1.01 (0.95, 1.08) | 0.66 |
| Aggressive | 1.01 (0.93, 1.10) | 0.80 |
| Pancreatic | 1.04 (0.92, 1.17) | 0.54 |
| Colorectal | 1.03 (0.97, 1.10) | 0.29 |
| Colon | 1.00 (0.94, 1.07) | 0.94 |
| Rectal | 1.04 (0.96, 1.13) | 0.31 |

Risk estimates based on multivariable Mendelian randomization. Risk estimates $p < 0.05$ are in bold. Abbreviations: CI=confidence interval; ER=estrogen receptor; OR=odds ratio.

## DISCUSSION

This study used both observational and MR methods to examine the relationship between cardiorespiratory fitness and incident cancer risk, providing the first evidence that higher fitness levels may reduce risks of breast cancer. In observational analysis only, we report additional inverse associations between VO_2_max scaled to total body mass and risks of colorectal and endometrial cancer. However, associations with all three cancer sites were attenuated after accounting for adiposity. Observational associations between cancer and VO_2_max scaled to fat-free mass were not statistically significant.

Previous observational analyses have reported inverse associations between fitness and colorectal and lung cancer, whereas we report that these relationships are null after accounting for adiposity^12–15^. Our results may differ from these previous studies due to differences in population sampling, fitness assessment, and fitness estimation approaches. For example, cycle ergometer-based fitness estimates may differ from treadmill-based estimates due to differences in load bearing and motion artefact^15, 16, 18^. The UK Biobank fitness test was also relatively light intensity, which enabled more participants to be assessed. Thus, our analysis likely characterizes a wider variety of lower-fitness individuals than previous studies which used more strenuous tests. Previous estimates using UK Biobank data had shorter duration of follow-up (median 5 years) and used fewer exercise test data, which will reduce the precision of risk estimates.

Previous MR studies based on up to five SNPs have reported inverse associations between genetically predicted physical activity levels and risks of breast, colorectal and aggressive prostate cancer^54, 55^. However, current estimates suggest that GWAS significant polymorphisms explain a very limited proportion of phenotypic physical activity (e.g. 0.06% for overall physical activity)^56^. The small number of SNPs increase the influence of possibly invalid variants within the instrument, and the instrument has a bidirectional association with BMI^56^.

Fitness is a trait that reflects both input from genetics and physical activity behaviours. The genetic instrument for fitness used in the present study likely encompasses both past and current levels as well as the capacity to participate in physical activity^2, 3^. This instrument explains 1.2% of the variation in observed fitness levels, increasing the reliability of risk estimates. Future work examining the relative importance of the different constituents of genetic fitness may help to clarify whether the null relationships that we report for fitness on colorectal and aggressive prostate cancer risk are indicative of the greater relative importance of physical activity behaviours or are partially reflective of the methodological limitations discussed above.

The role of adiposity in fitness is complex and not fully understood. Higher adiposity is associated with impaired physical performance, relating reduced muscle oxygen uptake, lower cardiac efficiency, neuromuscular dysfunction, and increased cancer risk^57–61^. Higher levels of physical activity are important for weight maintenance and increasing fitness, and higher fitness may reduce some of the harmful cardiometabolic effects of obesity^62^. Differences between the associations of fitness and cancer by scaling are likely driven by the different components of fitness, as VO_2_max_tbm_ has a strong inverse correlation with body size and adiposity^25^. However, the complex interplay of adiposity, fitness and cancer might mean that accounting for adiposity for models of cardiorespiratory fitness could lead to an over-adjustment of risk estimates, but these relationships are difficult to disentangle. Relationships between fitness and all-cause, cancer, and cardiovascular mortality outcomes has stronger evidence for independence of associations with adiposity^10, 41, 62, 63^. Future work with longer durations of follow-up will improve power to investigate whether there are differential risk associations by BMI classification.

These analyses have several strengths. This study is the first to use genetically instrumented fitness to evaluate possible causal relationships between fitness and cancer risk. The UK Biobank is the largest sample currently available with estimates of maximal cardiorespiratory fitness data, maximising power to assess associations across a broad range of cancer sites, the majority of which have not been previously investigated. Our independently validated novel framework to estimate fitness harmonised the UK Biobank test protocols and calibrated these data to a maximal exercise test to estimate VO₂max. This estimation framework also incorporated multiple heart rate measurements to reduce measurement error, with high temporal agreement (regression dilution ratio=0.79) over approximately a 2.8 year period for greater precision in risk estimates^42^. Further, the baseline assessment collected data across a wide range of lifestyle, medical and anthropometric factors, enabling thorough adjustment for possible confounders.

Our study has limitations. This analysis is not a randomised controlled trial and therefore we are not able to fully assess causality. Although MR may share some common design characteristics, we cannot exclude the possibility of genetic confounding or horizontal pleiotropy^64^. The genetic instrument for VO_2_max_tbm_ was not available for comparison with our observational analysis. The genetic instrument also included resting heart rate information; therefore, our results may be partially driven by genetic associations with resting heart rate.

Given the strong *a priori* evidence and mechanistic plausibility of associations between fitness and cancer risk we have not included correction for multiple testing^16–18^, however we cannot exclude the possibility of chance findings. The UK Biobank participants are predominantly of White European ancestry and are healthier than the underlying sampling population, therefore risk estimates may not be generalizable to some other populations, including “high-risk” participants who did not undergo the fitness assessment. The fitness test was also submaximal, which may increase measurement error, and previous studies have noted larger magnitudes of associations with health outcomes using maximal fitness tests^11, 15^.

In summary, we provide evidence that higher fitness levels may reduce risks of endometrial, colorectal, and breast cancer. The role of adiposity in mediating the relationship between fitness and cancer risk is not fully understood, and further research is needed to explore this complex relationship. Aiming to increase fitness, including via changes in body composition, may be an effective strategy to reduce risk of some cancer sites.

## DATA SHARING STATEMENT

The UK Biobank is an open-access resource and bona fide researchers can apply to use the UK Biobank dataset by registering and applying at http://ukbiobank.ac.uk/register-apply/. Further information is available from the corresponding author upon request. For genetic cancer data, summary GWAS statistics are publicly available for breast (https://bcac.ccge.medschl.cam.ac.uk/), endometrial (https://www.ebi.ac.uk/gwas/studies/GCST006464), ovarian (https://ocac.ccge.medschl.cam.ac.uk) and prostate (overall only) (http://practical.icr.ac.uk/).

Data for renal cell carcinoma and pancreatic cancer were accessed via dbGaP; Study Accession: phs000206.v3.p2 and phs000648.v1.p1; project reference 9314 (https://www.ncbi.nlm.nih.gov/gap/). Summary genetic data for aggressive prostate cancer (http://practical.icr.ac.uk/), lung cancer (https://ilcco.iarc.fr/) and colorectal cancer (https://research.fredhutch.org/peters/en/genetics-and-epidemiology-of-colorectal-cancer-consortium.html) are not currently publicly available but may be made available upon application, see respective websites for details.

## DATA ACCESS, RESPONSIBILITY AND ANALYSIS

TIG had full access to all the UK Biobank fitness data in the study and carried out the observational analysis. ELW had full access to all the genetic data in the study and carried out the MR analysis. They take responsibility for the integrity of the data and the accuracy of the data analysis.

## DISCLAIMER

The findings and conclusions in this article are those of the authors and do not necessarily represent the official position of the National Institutes of Health.

## Supporting information

NIH cover form

## Data Availability

The UK Biobank is an open-access resource and bona fide researchers can apply to use the UK Biobank dataset by registering and applying at http://ukbiobank.ac.uk/register-apply/. Further information is available from the corresponding author upon request. For genetic cancer data, summary GWAS statistics are publicly available for breast (https://bcac.ccge.medschl.cam.ac.uk/), endometrial (https://www.ebi.ac.uk/gwas/studies/GCST006464), ovarian (https://ocac.ccge.medschl.cam.ac.uk) and prostate (overall only) (http://practical.icr.ac.uk/). Data for renal cell carcinoma and pancreatic cancer were accessed via dbGaP; Study Accession: phs000206.v3.p2 and phs000648.v1.p1; project reference 9314 (https://www.ncbi.nlm.nih.gov/gap/). Summary genetic data for aggressive prostate cancer (http://practical.icr.ac.uk/), lung cancer (https://ilcco.iarc.fr/) and colorectal cancer (https://research.fredhutch.org/peters/en/genetics-and-epidemiology-of-colorectal-cancer-consortium.html) are not currently publicly available but may be made available upon application, see respective websites for details.

## ACKNOWLEDGEMENTS

This research has been conducted using the UK Biobank Resource under application number #408 and #9905. The authors have no conflicts of interest to disclose. This work utilized the computational resources of the NIH HPC Biowulf cluster. (http://hpc.nih.gov).

We thank all participants, researchers and support staff who made the study possible. We additionally would like to thank BCAC, ECAC, GECCO, ILCCO, OCAC, PanScan, PanC4 and the PRACTICAL consortia for their invaluable contributions to the genetic analysis. Genetic consortium specific acknowledgements and funding information are available from Appendix 2.

## REFERENCES

1. Moore SC, Lee IM, Weiderpass E, et al. Association of leisure-time physical activity with risk of 26 types of cancer in 1.44 Million adults. JAMA Intern Med. 2016;176(6):816–825. doi:10.1001/jamainternmed.2016.1548

2. Sarzynski MA, Ghosh S, Bouchard C. Genomic and transcriptomic predictors of response levels to endurance exercise training. J Physiol. 2017;595(9):2931–2939. doi:10.1113/JP272559

3. Kim DS, Wheeler MT, Ashley EA. The genetics of human performance. Nat Rev Genet. 2022;23(1):40–54. doi:10.1038/s41576-021-00400-5

4. Bouchard C, An P, Rice T, et al. Familial aggregation of VO2max response to exercise training: Results from the HERITAGE Family Study. J Appl Physiol. 1999;87(3):1003–1008. doi:10.1152/jappl.1999.87.3.1003

5. Al-Mallah MH, Sakr S, Al-Qunaibet A. Cardiorespiratory fitness and cardiovascular disease prevention: An update. Curr Atheroscler Rep. 2018;20(1):1. doi:10.1007/s11883-018-0711-4

6. Kaze AD, Agoons DD, Santhanam P, Erqou S, Ahima RS, Echouffo-Tcheugui JB. Correlates of cardiorespiratory fitness among overweight or obese individuals with type 2 diabetes. BMJ Open Diabetes Res Care. 2022;10(1):e002446. doi:10.1136/bmjdrc-2021- 002446

7. LaMonte MJ, Eisenman PA, Adams TD, Shultz BB, Ainsworth BE, Yanowitz FG. Cardiorespiratory fitness and coronary heart disease risk factors: the LDS Hospital Fitness Institute cohort. Circulation. 2000;102(14):1623–1628. doi:10.1161/01.cir.102.14.1623

8. Arsenault BJ, Lachance D, Lemieux I, et al. Visceral Adipose Tissue Accumulation, Cardiorespiratory Fitness, and Features of the Metabolic Syndrome. Arch Intern Med. 2007;167(14):1518–1525. doi:10.1001/archinte.167.14.1518

9. Han M, Qie R, Shi X, et al. Cardiorespiratory fitness and mortality from all causes, cardiovascular disease and cancer: Dose–response meta-analysis of cohort studies. Br J Sports Med. Published online January 12, 2022. doi:10.1136/bjsports-2021-104876

10. Schmid D, Leitzmann MF. Cardiorespiratory fitness as predictor of cancer mortality: a systematic review and meta-analysis. Ann Oncol. 2015;26(2):272–278. doi:10.1093/annonc/mdu250

11. Laukkanen JA, Isiozor NM, Kunutsor SK. Objectively Assessed Cardiorespiratory Fitness and All-Cause Mortality Risk: An Updated Meta-analysis of 37 Cohort Studies Involving 2,258,029 Participants. Mayo Clin Proc. 2022;97(6):1054–1073. doi:10.1016/j.mayocp.2022.02.029

12. Marshall CH, Al-Mallah MH, Dardari Z, et al. Cardiorespiratory fitness and incident lung and colorectal cancer in men and women: Results from the Henry Ford Exercise Testing (FIT) cohort. Cancer. 2019;125(15):2594–2601. doi:10.1002/cncr.32085

13. Pozuelo-Carrascosa DP, Alvarez-Bueno C, Cavero-Redondo I, Morais S, Lee IM, Martínez-Vizcaíno V. Cardiorespiratory fitness and site-specific risk of cancer in men: A systematic review and meta-analysis. Eur J Cancer. 2019;113:58–68. doi:10.1016/j.ejca.2019.03.008

14. Steell L, Ho FK, Sillars A, et al. Dose-response associations of cardiorespiratory fitness with all-cause mortality and incidence and mortality of cancer and cardiovascular and respiratory diseases: the UK Biobank cohort study. Br J Sports Med. 2019;53(21):1371–1378. doi:10.1136/bjsports-2018-099093

15. Lakoski SG, Willis BL, Barlow CE, et al. Midlife Cardiorespiratory Fitness, Incident Cancer, and Survival After Cancer in Men: The Cooper Center Longitudinal Study. JAMA Oncol. 2015;1(2):231–237. doi:10.1001/jamaoncol.2015.0226

16. Vainshelboim B, Müller J, Lima RM, et al. Cardiorespiratory fitness and cancer incidence in men. Ann Epidemiol. 2017;27(7):442–447. doi:10.1016/j.annepidem.2017.06.003

17. Kunutsor SK, Voutilainen A, Laukkanen JA. Cardiorespiratory fitness is not associated with reduced risk of prostate cancer: A cohort study and review of the literature. Eur J Clin Invest. 2021;51(8):e13545. doi:10.1111/eci.13545

18. Byun W, Sui X, Hébert JR, et al. Cardiorespiratory fitness and risk of prostate cancer: findings from the Aerobics Center Longitudinal Study. Cancer Epidemiol. 2011;35(1):59–65. doi:10.1016/j.canep.2010.07.013

19. Cai L, Gonzales T, Wheeler E, et al. Causal associations between cardiorespiratory fitness and type 2 diabetes. Nat Commun [Accepted]

20. Lawlor DA, Tilling K, Davey Smith G. Triangulation in aetiological epidemiology. Int J Epidemiol. 2016;45(6):1866–1886. doi:10.1093/ije/dyw314

21. UK Biobank: Protocol for a large-scale prospective epidemiological resource. Published online March 21, 2007. https://www.ukbiobank.ac.uk/media/gnkeyh2q/study-rationale.pdf

22. UK Biobank cardio assessment manual: Version 1.0. Published online April 5, 2011. http://biobank.ctsu.ox.ac.uk/crystal/docs/Cardio.pdf

23. Imboden MT, Kaminsky LA, Peterman JE, et al. Cardiorespiratory fitness normalized to fat-free mass and mortality risk. Med Sci Sports Exerc. 2020;52(7):1532–1537. doi:10.1249/MSS.0000000000002289

24. Osman AF, Mehra MR, Lavie CJ, Nunez E, Milani RV. The incremental prognostic importance of body fat adjusted peak oxygen consumption in chronic heart failure. J Am Coll Cardiol. 2000;36(7):2126–2131. doi:10.1016/S0735-1097(00)00985-2

25. Zhou N. Assessment of aerobic exercise capacity in obesity, which expression of oxygen uptake is the best? Sports Med Health Sci. 2021;3(3):138–147. doi:10.1016/j.smhs.2021.01.001

26. Bowden J, Spiller W, Del Greco M F, et al. Improving the visualization, interpretation and analysis of two-sample summary data Mendelian randomization via the Radial plot and Radial regression. Int J Epidemiol. 2018;47(4):1264–1278. doi:10.1093/ije/dyy101

27. Kokkinos P, Myers J, Franklin B, Narayan P, Lavie CJ, Faselis C. Cardiorespiratory fitness and health outcomes: A call to standardize fitness categories. Mayo Clin Proc. 2018;93(3):333–336. doi:10.1016/j.mayocp.2017.10.011

28. Haycock PC, Burgess S, Wade KH, Bowden J, Relton C, Davey Smith G. Best (but oft-forgotten) practices: the design, analysis, and interpretation of Mendelian randomization studies1. Am J Clin Nutr. 2016;103(4):965–978. doi:10.3945/ajcn.115.118216

29. Michailidou K, Lindström S, Dennis J, et al. Association analysis identifies 65 new breast cancer risk loci. Nature. 2017;551(7678):92–94. doi:10.1038/nature24284

30. Zhang H, Ahearn TU, Lecarpentier J, et al. Genome-wide association study identifies 32 novel breast cancer susceptibility loci from overall and subtype-specific analyses. Nat Genet. 2020;52(6):572–581. doi:10.1038/s41588-020-0609-2

31. Schumacher FR, Al Olama AA, Berndt SI, et al. Association analyses of more than 140,000 men identify 63 new prostate cancer susceptibility loci. Nat Genet. 2018;50(7):928–936. doi:10.1038/s41588-018-0142-8

32. O’Mara TA, Glubb DM, Amant F, et al. Identification of nine new susceptibility loci for endometrial cancer. Nat Commun. 2018;9(1):3166. doi:10.1038/s41467-018-05427-7

33. Phelan CM, Kuchenbaecker KB, Tyrer JP, et al. Identification of 12 new susceptibility loci for different histotypes of epithelial ovarian cancer. Nat Genet. 2017;49(5):680–691. doi:10.1038/ng.3826

34. McKay JD, Hung RJ, Han Y, et al. Large-scale association analysis identifies new lung cancer susceptibility loci and heterogeneity in genetic susceptibility across histological subtypes. Nat Genet. 2017;49(7):1126–1132. doi:10.1038/ng.3892

35. Huyghe JR, Bien SA, Harrison TA, et al. Discovery of common and rare genetic risk variants for colorectal cancer. Nat Genet. 2019;51(1):76–87. doi:10.1038/s41588-018-0286-6

36. Huyghe JR, Harrison TA, Bien SA, et al. Genetic architectures of proximal and distal colorectal cancer are partly distinct. Gut. 2021;70(7):1325–1334. doi:10.1136/gutjnl-2020-321534

37. Scelo G, Purdue MP, Brown KM, et al. Genome-wide association study identifies multiple risk loci for renal cell carcinoma. Nat Commun. 2017;8(1):15724. doi:10.1038/ncomms15724

38. Childs EJ, Mocci E, Campa D, et al. Common variation at 2p13.3, 3q29, 7p13 and 17q25.1 associated with susceptibility to pancreatic cancer. Nat Genet. 2015;47(8):911–916. doi:10.1038/ng.3341

39. Petersen GM, Amundadottir L, Fuchs CS, et al. A genome-wide association study identifies pancreatic cancer susceptibility loci on chromosomes 13q22.1, 1q32.1 and 5p15.33. Nat Genet. 2010;42(3):224–228. doi:10.1038/ng.522

40. Amundadottir L, Kraft P, Stolzenberg-Solomon RZ, et al. Genome-wide association study identifies variants in the ABO locus associated with susceptibility to pancreatic cancer. Nat Genet. 2009;41(9):986–990. doi:10.1038/ng.429

41. Farrell SW, Cortese GM, LaMonte MJ, Blair SN. Cardiorespiratory fitness, different measures of adiposity, and cancer mortality in men. Obesity. 2007;15(12):3140–3149. doi:10.1038/oby.2007.374

42. Gonzales TI, Westgate K, Strain T, et al. Cardiorespiratory fitness assessment using risk-stratified exercise testing and dose-response relationships with disease outcomes. Sci Rep. 2021;11(1):15315. doi:10.1038/s41598-021-94768-3

43. Keogh RH, White IR. A toolkit for measurement error correction, with a focus on nutritional epidemiology. Stat Med. 2014;33(12):2137–2155. doi:10.1002/sim.6095

44. Burgess S, Bowden J. Integrating summarized data from multiple genetic variants in Mendelian randomization: Bias and coverage properties of inverse-variance weighted methods. Published online November 27, 2015. Accessed May 13, 2022. http://arxiv.org/abs/1512.04486

45. Burgess S, Thompson SG, CRP CHD Genetics Collaboration. Avoiding bias from weak instruments in Mendelian randomization studies. Int J Epidemiol. 2011;40(3):755–764. doi:10.1093/ije/dyr036

46. Burgess S, Bowden J, Fall T, Ingelsson E, Thompson SG. Sensitivity analyses for robust causal inference from Mendelian randomization analyses with multiple genetic variants. Epidemiol Camb Mass. 2017;28(1):30–42. doi:10.1097/EDE.0000000000000559

47. Kamat MA, Blackshaw JA, Young R, et al. PhenoScanner V2: An expanded tool for searching human genotype-phenotype associations. Bioinforma Oxf Engl. 2019;35(22):4851–4853. doi:10.1093/bioinformatics/btz469

48. Verbanck M, Chen CY, Neale B, Do R. Detection of widespread horizontal pleiotropy in causal relationships inferred from Mendelian randomization between complex traits and diseases. Nat Genet. 2018;50(5):693–698. doi:10.1038/s41588-018-0099-7

49. Bowden J, Davey Smith G, Haycock PC, Burgess S. Consistent Estimation in Mendelian Randomization with Some Invalid Instruments Using a Weighted Median Estimator. Genet Epidemiol. 2016;40(4):304–314. doi:10.1002/gepi.21965

50. UK Biobank. Neale lab. Accessed August 2, 2022. http://www.nealelab.is/uk-biobank

51. Hemani G, Zheng J, Elsworth B, et al. The MR-Base platform supports systematic causal inference across the human phenome. eLife. 2018;7:e34408. doi:10.7554/eLife.34408

52. Yavorska OO, Burgess S. MendelianRandomization: an R package for performing Mendelian randomization analyses using summarized data. Int J Epidemiol. 2017;46(6):1734–1739. doi:10.1093/ije/dyx034

53. Bowden J, Del Greco M F, Minelli C, Davey Smith G, Sheehan NA, Thompson JR. Assessing the suitability of summary data for two-sample Mendelian randomization analyses using MR-Egger regression: the role of the I2 statistic. Int J Epidemiol. 2016;45(6):1961–1974. doi:10.1093/ije/dyw220

54. Kazmi N, Haycock P, Tsilidis K, et al. Appraising causal relationships of dietary, nutritional and physical-activity exposures with overall and aggressive prostate cancer: two-sample Mendelian-randomization study based on 79 148 prostate-cancer cases and 61 106 controls. Int J Epidemiol. 2020;49(2):587–596. doi:10.1093/ije/dyz235

55. Papadimitriou N, Dimou N, Tsilidis KK, et al. Physical activity and risks of breast and colorectal cancer: a Mendelian randomisation analysis. Nat Commun. 2020;11(1):597. doi:10.1038/s41467-020-14389-8

56. Doherty A, Smith-Byrne K, Ferreira T, et al. GWAS identifies 14 loci for device-measured physical activity and sleep duration. Nat Commun. 2018;9(1):5257. doi:10.1038/s41467-018-07743-4

57. Wearing SC, Hennig EM, Byrne NM, Steele JR, Hills AP. The impact of childhood obesity on musculoskeletal form. Obes Rev Off J Int Assoc Study Obes. 2006;7(2):209–218. doi:10.1111/j.1467-789X.2006.00216.x

58. Hung TH, Liao PA, Chang HH, Wang JH, Wu MC. Examining the relationship between cardiorespiratory fitness and body weight status: empirical evidence from a population-based survey of adults in Taiwan. ScientificWorldJournal. 2014;2014:463736. doi:10.1155/2014/463736

59. Tanner CJ, Barakat HA, Dohm GL, et al. Muscle fiber type is associated with obesity and weight loss. Am J Physiol Endocrinol Metab. 2002;282(6):E1191–1196. doi:10.1152/ajpendo.00416.2001

60. Norman AC, Drinkard B, McDuffie JR, Ghorbani S, Yanoff LB, Yanovski JA. Influence of excess adiposity on exercise fitness and performance in overweight children and adolescents. Pediatrics. 2005;115(6):e690–696. doi:10.1542/peds.2004-1543

61. Larsson SC, Spyrou N, Mantzoros CS. Body fatness associations with cancer: evidence from recent epidemiological studies and future directions. Metabolism. 2022;137:155326. doi:10.1016/j.metabol.2022.155326

62. Barry VW, Baruth M, Beets MW, Durstine JL, Liu J, Blair SN. Fitness vs. fatness on all-cause mortality: a meta-analysis. Prog Cardiovasc Dis. 2014;56(4):382–390. doi:10.1016/j.pcad.2013.09.002

63. Sui X, LaMonte MJ, Laditka JN, et al. Cardiorespiratory Fitness and Adiposity as Mortality Predictors in Older Adults. JAMA J Am Med Assoc. 2007;298(21):2507–2516. doi:10.1001/jama.298.21.2507

64. Davies NM, Holmes MV, Smith GD. Reading Mendelian randomisation studies: a guide, glossary, and checklist for clinicians. BMJ. 2018;362:k601. doi:10.1136/bmj.k601

