## Supplementary Materials for "Observational and genetic associations between cardiorespiratory fitness and cancer: a UK Biobank and international consortia study"

+ Joint senior authors


### Supplementary Materials

### Supplemental Methods

*UK Biobank test description*

Participants were assigned an exercise risk level (minimal, small, medium, and high) according to responses on a modified Rose-Angina Questionnaire and measured blood pressure. Those with "minimal" and "small" risk completed an incremental test of varying intensity, those with "medium" risk completed a low-intensity steady-state test, and those with "high" risk did not complete an exercise test. Incremental tests had two sequential phases: 2 minutes of cycling at a fixed work rate (30 watts for women, 40 watts for men) and 4 minutes of cycling while work rate increased in small increments every few seconds to a target work rate. The target work rate was defined to be at most 50% of the participant's estimated maximal work, which was computed using participant age, sex, height, weight, resting heart rate, and exercise risk level.[1] Steady-state tests consisted of 6 minutes of cycling only. All tests concluded with a 1 minute recovery period. Cycling was performed on an electromagnetically-braked stationary bike (eBike ergometer, GE) at a 60 revolutions per minute cadence. Heart rate response to exercise was recorded continuously using 4-lead electrocardiography (Cardiosoft) on the forearms.

*Maximal oxygen consumption estimation*

Maximal oxygen consumption ($VO_2$ max; ml $O_2 \cdot min^{-1} \cdot kg^{-1}$) was estimated from the linear relationship between exercise heart rate response and cycling work rate using a multilevel estimation framework (Supplementary Figure 1). This estimation method has been validated against direct $VO_2$max measurements, demonstrating moderate correlation (Pearson's r: 0.70) and zero overall mean bias between estimated and directly measured $VO_2$max; differential bias by sex was minimal. In the present analysis, $VO_2$ max was estimated in two ways: scaled by total-body mass ($VO_2max_{tbm}$ [3.5 ml $O_2 \cdot min^{-1} \cdot kg^{-1}$ total-body mass=1 MET]) and scaled by fat-free mass ($VO_2max_{ffm}$). $VO_2max_{ffm}$ was computed by multiplying $VO_2max_{tbm}$ by total-body mass and then dividing by fat-free mass.

*Cancer site definitions*

Cancer sites were defined using the International Classification of Diseases Tenth revision codes [ICD-10] as follows: breast (C50), prostate (C61), colorectal (C18-20, C26), colon (C18, C26), rectum (C19-20), lung (C34), endometrial (C54-55) and ovarian (C56), excluding the ICD-0-3 histologies (9050-9055, 9140, 9590-9992). Cancer sites for inclusion for survival analyses were based on the minimum number of cases (n>100), and the availability of large-scale genetic consortia. Person-years were calculated from the date of recruitment to the date of the first cancer registration (excluding non-melanoma skin cancer [ICD-10 C44]), death or censoring date, whichever occurred first.

*Cox regression model adjustment description*

Cox regression models were adjusted for sex, self-reported racial/ethnic group (Asian or Asian British, Black or Black British, Mixed, Other, White), Townsend index of deprivation, education (no qualifications, any other qualification, degree level or above), employment status (unemployed, employed, retired), smoking status (never, current, previous), alcohol consumption (never or previous, 1 or 2 times per week, 3 or more times per week), red and processed meat consumption (average days per week derived from questions on the frequency of processed, beef, lamb and pork intake), fish consumption (never, less than once per week, at least once per week), fruit and vegetable consumption (a score of '0–4' was computed from self-reported intake frequency of raw vegetables, cooked vegetables, fresh fruit, and dried fruit), salt consumption (never or rarely, sometimes, usually or always), diabetes status (binary variable set to '1' if participants self-reported a diabetes diagnosis or current insulin therapy; '0' otherwise), hypertension (binary variable set to '1' if one of the following were observed: measured systolic blood pressure greater or equal to 140, measured diastolic blood pressure greater or equal to 90, or self-reported use of blood pressure medication; '0' otherwise), and other medication use (separate binary variables set to '1' if reported use of each of the following: beta blockers, calcium channel blockers, angiotensin-converting enzyme inhibitors, diuretics, bronchodilators, lipid-lowering agents, iron deficiency agents, non-steroidal anti-inflammatory drugs, metformin; '0' otherwise). Female reproductive cancers (breast, endometrial, and ovarian) were additionally adjusted for age at menarche, age at menopause, parity, hormone replacement therapy usage, and oral contraceptives. Multivariate imputation by chained equations was used to impute missing covariate values. Adiposity may partially mediate and confound the relationship between fitness and cancer risk. We therefore evaluated the role of adiposity in fitness-to-cancer associations by running all models with and without adjustment for either continuous BMI (for models with $VO_2max$ scaled by total-body mass) or fat mass (for models with $VO_2max$ scaled by fat-free mass).

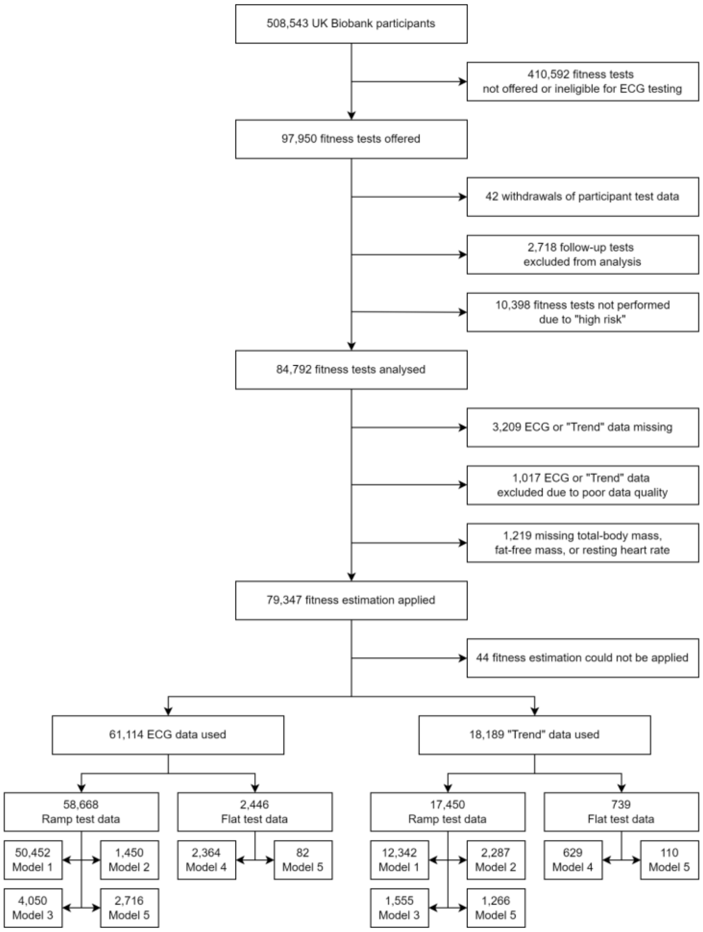


**Supplementary Figure 1**

Flow diagram representing the number of participants included for the estimation of cardiorespiratory fitness and allocation of multilevel framework estimation models:

Model 1: based on ramp and recovery phase. Covariates: $HR_{max}$, $HR_{rest}$, $HR_{rec45}$, $b_0$, $b_1$, $RR^{0.5}$, sex
Model 2: based on recovery phase. Covariates: $HR_{max}$, $HR_{rest}$, $HR_{rec45}$, $RR^{0.5}$, sex, $HR_{rec0}$
Model 3: based on ramp phase. Covariates: $HR_{max}$, $HR_{rest}$, $b_0$, $b_1$, $RR^{0.5}$, sex
Model 4: based on flat and recovery phases. Covariates: $HR_{max}$, $HR_{rec45}$, sex, $HR_{flat}$
Model 5: based on flat phase. Covariates: $HR_{max}$, $HR_{rest}$, sex, $HR_{flat}$

"Trend" data represents instantaneous HR values computed using a proprietary algorithm in the software used to record data (Cardiosoft). For some test sessions, this is the only data available for analysis because raw ECG data were missing. Models were allocated on the basis of data availability after quality controls, where ECG data were preferentially used in analysis when available.

The models described above were used to estimate participant's maximal achieved work rate that would be achieved if a steady-state test had been conducted. Estimated work rate values were converted to $VO_2$ values using the American College of Sports Medicine metabolic equation for cycle ergometry:

$$VO_2max = 1.8 \cdot 6.12 \cdot \frac{Predicted\ work\ rate}{Participant\ weight} + 7$$

Further details of quality control, model allocation and coefficients are available from Gonzales *et al.*, 2021. [2]

Abbreviations: $b_0$= intercept from ramp phase linear regression model; $b_1$=slope from ramp phase linear regression model; HR= heart rate; $HR_{flat}$= median HR computed for the flat phase; $HR_{max}$= maximum heart rate (either age-predicted or directly measured); $HR_{rec0}$= heart rate 0 seconds post exercise; $HR_{rec45}$= recovery heart rate 45 seconds post exercise; $HR_{rest}$= resting heart rate; $RR^{0.5}$=square root of the test ramp rate

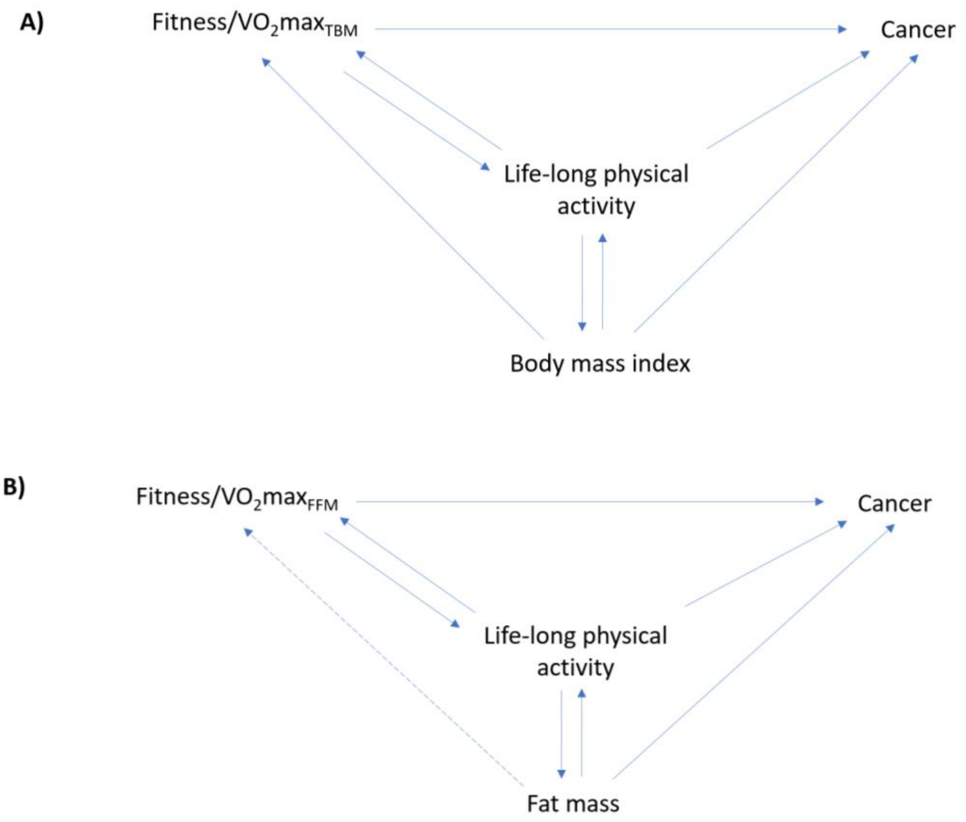


**Supplementary Figure 2**

Conceptual model of hypothesized relationships between fitness, lifelong physical activity (unobservable), adiposity, and cancer by fitness scaling
Dashed line between fat mass and $VO_2max_{FFM}$ depicts the weaker relationship with adiposity in comparison with $VO_2max_{TBM}$
Abbreviations: FFM=fat free mass; TBM=total body mass

### Supplementary Figure 3

HR and 95% CI for nonlinear associations (cubic splines, Cox regression) between cardiorespiratory fitness (per ml $O_2 \cdot min^{-1} \cdot kg^{-1}$ total-body mass and fat-free mass) and incident cancer. Knots were placed at the 33rd and 67th percentile of the fitness distribution. The reference fitness value was set to the mean value for the analytic sample. Age was the underlying time variable and models were adjusted for sex, ethnic group, Townsend deprivation index, education, employment status, smoking status, alcohol consumption, red and processed meat consumption, fish consumption, fruit and vegetable consumption, salt consumption, diabetes status, hypertension, medication use (beta blockers, calcium channel blockers, angiotensin-converting enzyme inhibitors, diuretics, bronchodilators, lipid-lowering agents, iron deficiency agents, non-steroid anti-inflammatory drugs, metformin). Female reproductive cancers (breast, endometrial, and ovarian) were additionally adjusted for age at menarche, age at menopause, parity, hormone replacement therapy usage, and oral contraceptives. Associations with and without adjustment for either continuous BMI (for models with $VO_2max$ scaled by total-body mass) or fat mass (for models with $VO_2max$ scaled by fat-free mass). Pooled reference values: $VO_2max_{tbm}$: 28.5 ml $O_2 \cdot min^{-1} \cdot kg^{-1}$, $VO_2max_{ffm}$: 41.0 ml $O_2 \cdot min^{-1} \cdot kg^{-1}$; Female-specific cancer reference values: $VO_2max_{tbm}$: 25.4 ml $O_2 \cdot min^{-1} \cdot kg^{-1}$, $VO_2max_{ffm}$: 39.6 ml $O_2 \cdot min^{-1} \cdot kg^{-1}$; Male-specific cancer reference values: $VO_2max_{tbm}$: 32.0 ml $O_2 \cdot min^{-1} \cdot kg^{-1}$, $VO_2max_{ffm}$: 42.6 ml $O_2 \cdot min^{-1} \cdot kg^{-1}$; Non-smoker reference values: $VO_2max_{tbm}$: 28.4 ml $O_2 \cdot min^{-1} \cdot kg^{-1}$, $VO_2max_{ffm}$: 40.9 ml $O_2 \cdot min^{-1} \cdot kg^{-1}$)

**HR per ml $O_2 \cdot min^{-1} \cdot kg^{-1}$ total-body mass**

**HR per ml $O_2 \cdot min^{-1} \cdot kg^{-1}$ fat-free mass**

**Maximally adjusted without BMI adjustment**

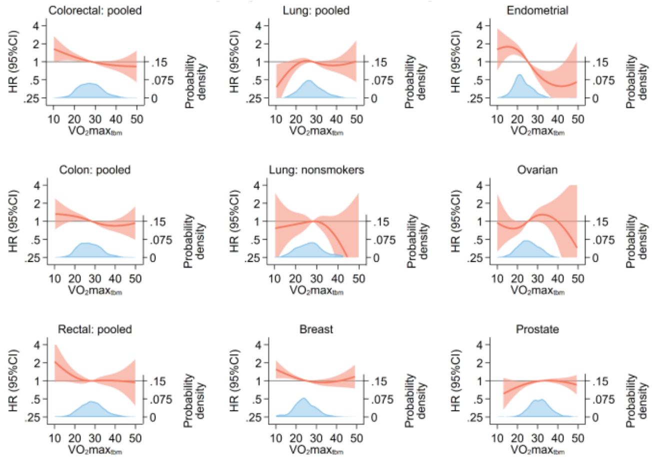


**Maximally adjusted without fat mass adjustment**

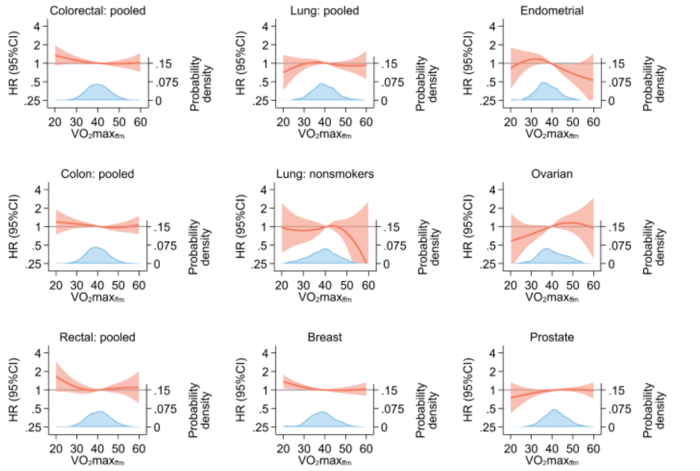


**Maximally adjusted with BMI adjustment**

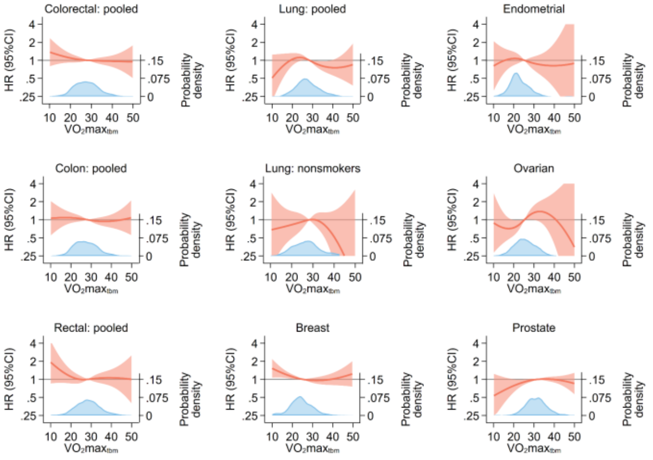


**Maximally adjusted with fat mass adjustment**

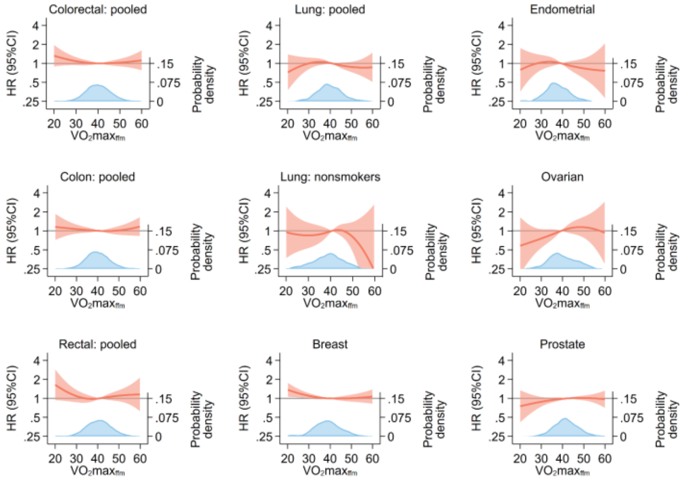

**Supplementary Figure 4**

HR and 95% CI for sex-stratified and nonlinear associations (cubic splines, Cox regression) between cardiorespiratory fitness (per ml $O_2 \cdot min^{-1} \cdot kg^{-1}$ total-body mass and fat-free mass) and incident cancer. Knots were placed at the 33rd and 67th percentile of the fitness distribution. The reference fitness value was set to the mean value for the analytic sample. Age was the underlying time variable and models were adjusted for sex, ethnic group, Townsend deprivation index, education, employment status, smoking status, alcohol consumption, red and processed meat consumption, fish consumption, fruit and vegetable consumption, salt consumption, diabetes status, hypertension, medication use (beta blockers, calcium channel blockers, angiotensin-converting enzyme inhibitors, diuretics, bronchodilators, lipid-lowering agents, iron deficiency agents, non-steroid anti-inflammatory drugs, metformin). Female reproductive cancers (breast, endometrial, and ovarian) were additionally adjusted for age at menarche, age at menopause, parity, hormone replacement therapy usage, and oral contraceptives. Associations with and without adjustment for either continuous BMI (for models with $VO_2max$ scaled by total-body mass) or fat mass (for models with $VO_2max$ scaled by fat-free mass). Women reference values: $VO_2max_{tbm}$: 25.4 ml $O_2 \cdot min^{-1} \cdot kg^{-1}$, $VO_2max_{ffm}$: 39.6 ml $O_2 \cdot min^{-1} \cdot kg^{-1}$; men reference values: $VO_2max_{tbm}$: 32.0 ml $O_2 \cdot min^{-1} \cdot kg^{-1}$, $VO_2max_{ffm}$: 42.6 ml $O_2 \cdot min^{-1} \cdot kg^{-1}$).

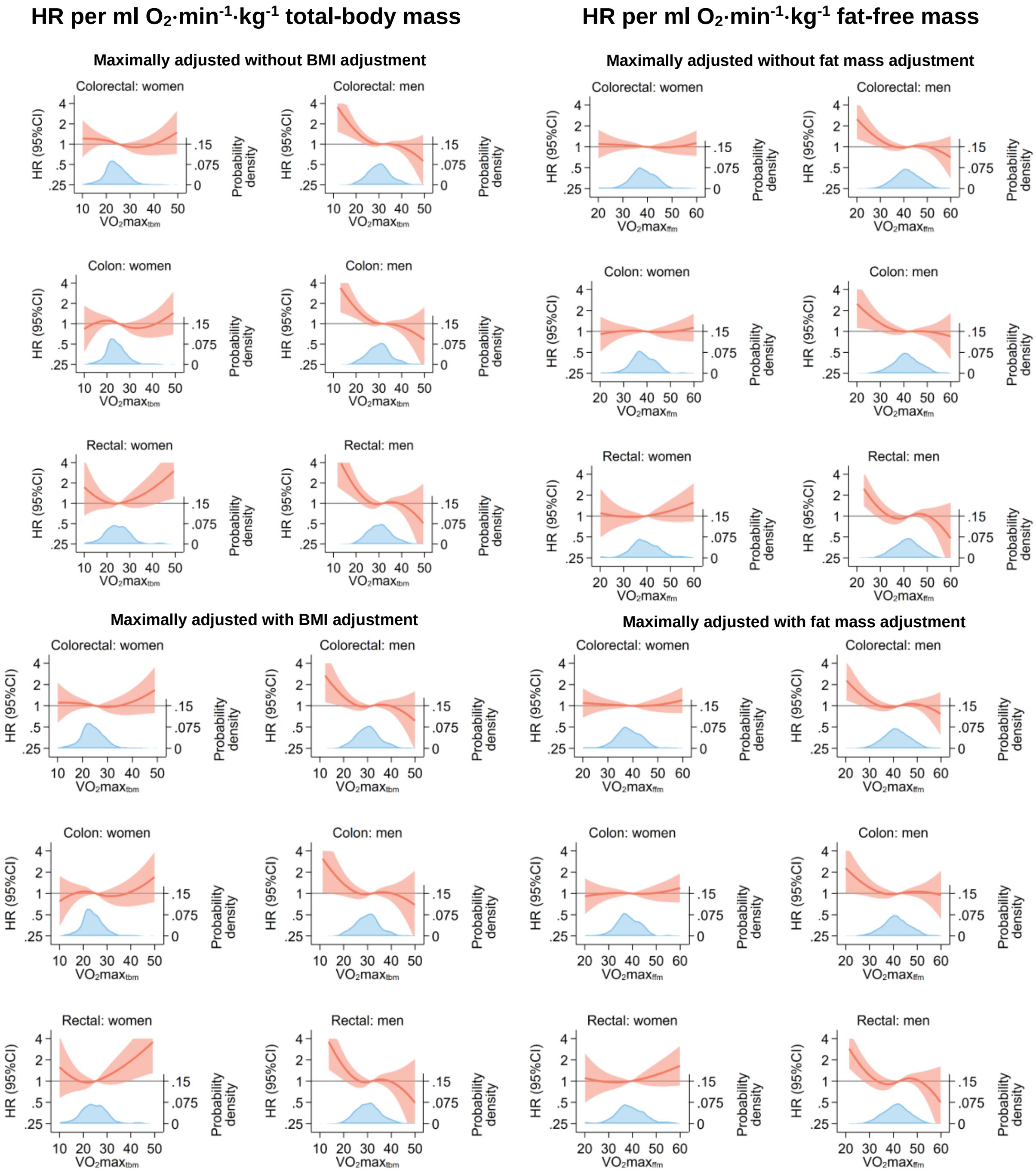

**Supplementary Figure 5**

HR and 95% CI for associations between cardiorespiratory fitness and incident cancer, both unadjusted for measurement error (i.e undiluted) and adjusted for measurement error (i.e. diluted). HRs and 95% CIs estimated using Cox regression models adjusted for age, sex, self-reported racial/ethnic group, Townsend index of deprivation, education, employment status, smoking status, alcohol consumption, red and processed meat consumption, fish consumption, fruit and vegetable consumption, salt consumption, diabetes status, hypertension, medication use (beta blockers, calcium channel blockers, ACE inhibitors, diuretics, bronchodilators, lipid-lowering agents, iron deficiency agents, non-steroidal anti-inflammatory drugs, metformin). Female reproductive cancers (breast, endometrial, and ovarian) were additionally adjusted for age at menarche, age at menopause, parity, hormone replacement therapy usage, and oral contraceptives. Regression dilution coefficient and standard deviation were 0.788 and 0.0118, respectively.

**HR per 3.5 ml $O_2 \cdot min^{-1} \cdot kg$ total-body mass**

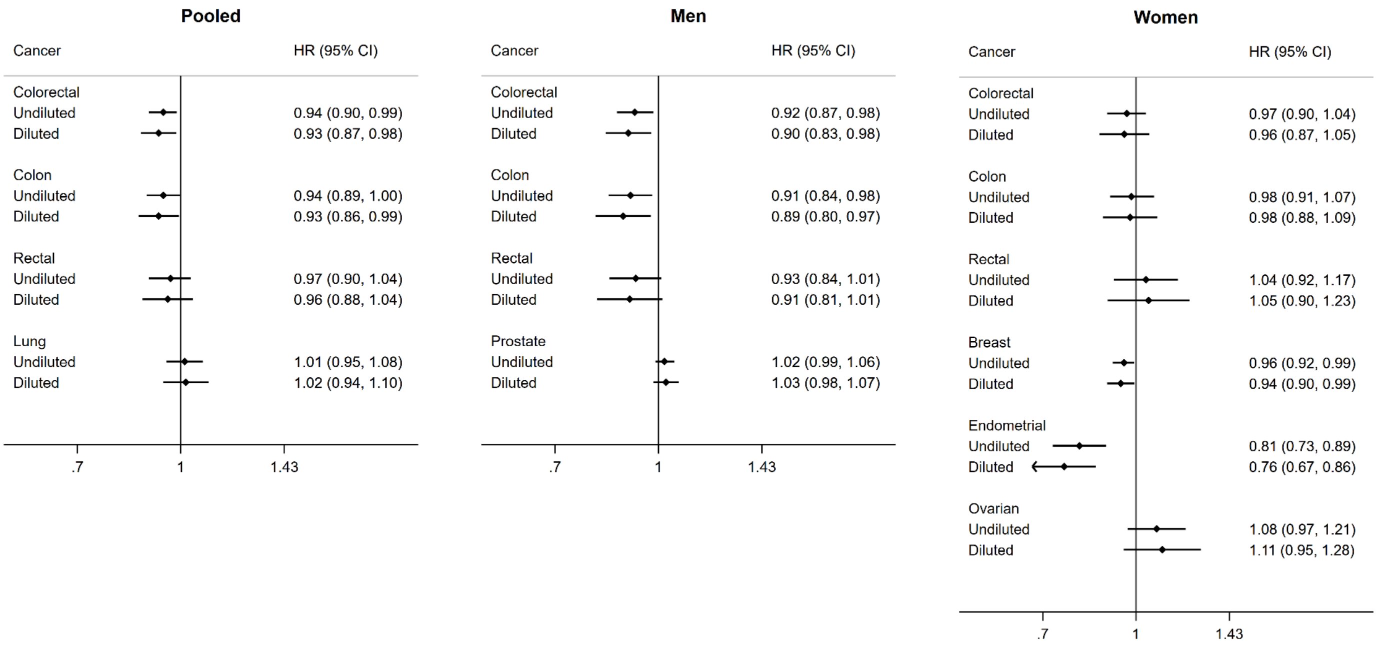


**HR per 5.0 ml $O_2 \cdot min^{-1} \cdot kg$ fat-free mass**

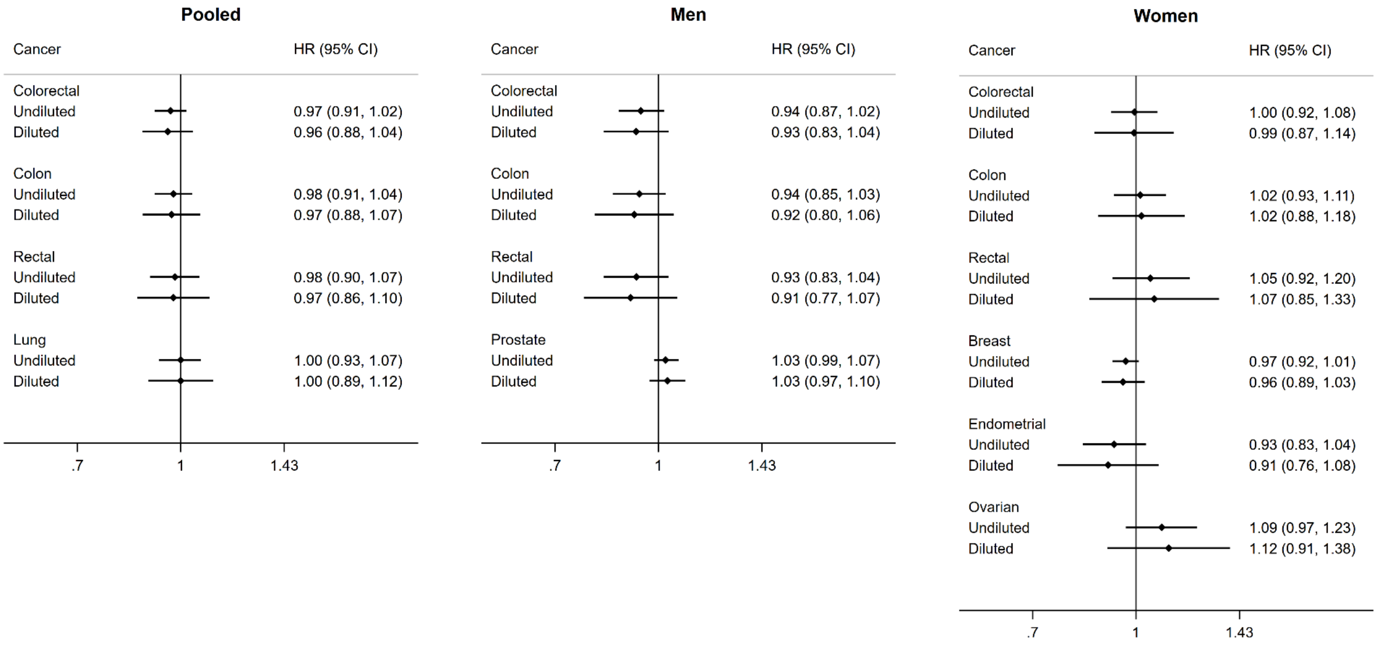

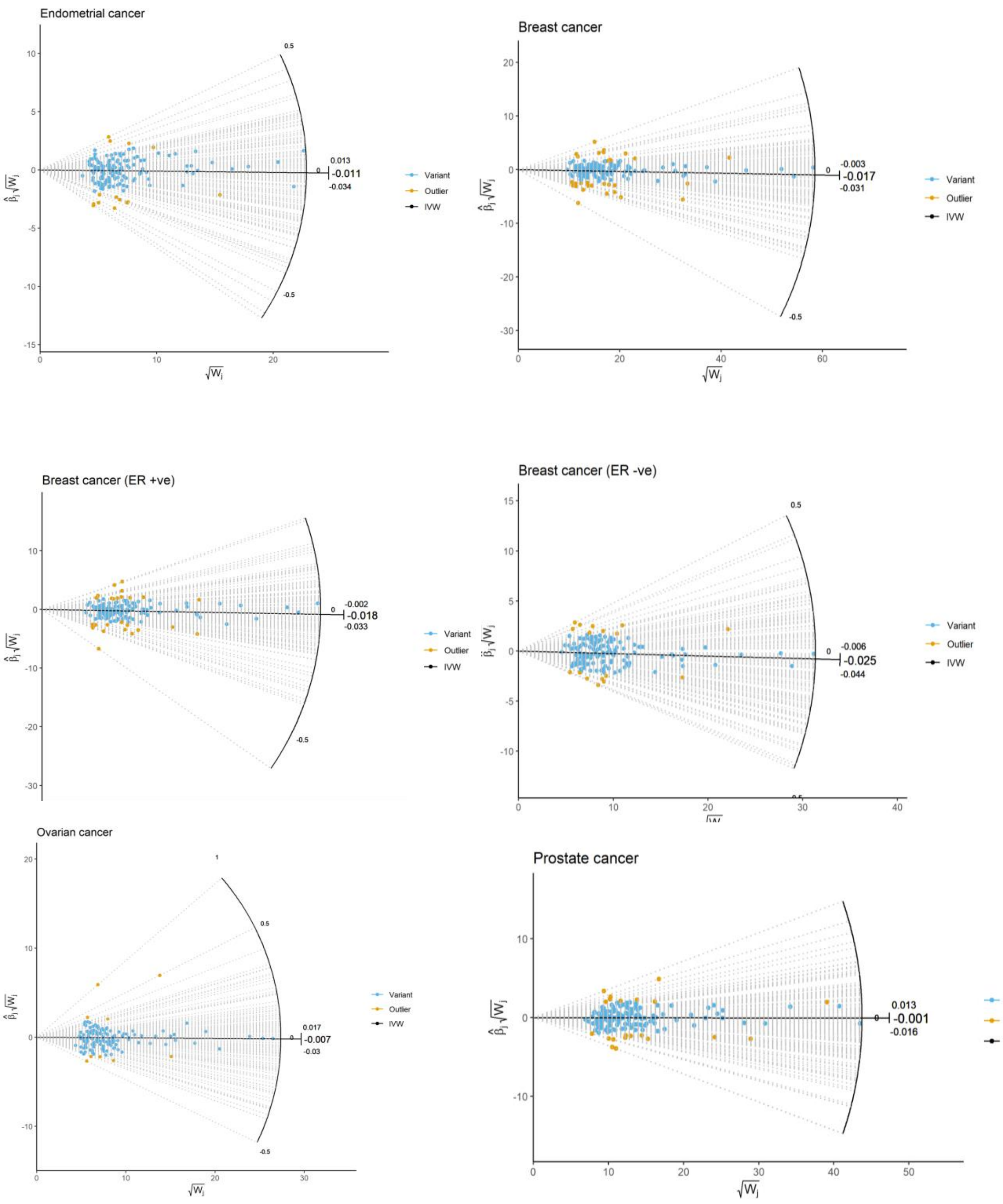


**Supplementary Figure 6: Radial plots of genetically predicted fitness with cancer outcomes**

Radial plots to visualize outlier SNPs in Mendelian randomization analysis. The radial curve shows the ratio estimate for each genetic variant and the IVW estimate. The lines join each data point back to the origin. Plots were produced using the RadialMR package[3].

Abbreviations: IVW= inverse variance weighted; SNP=single nucleotide polymorphism.

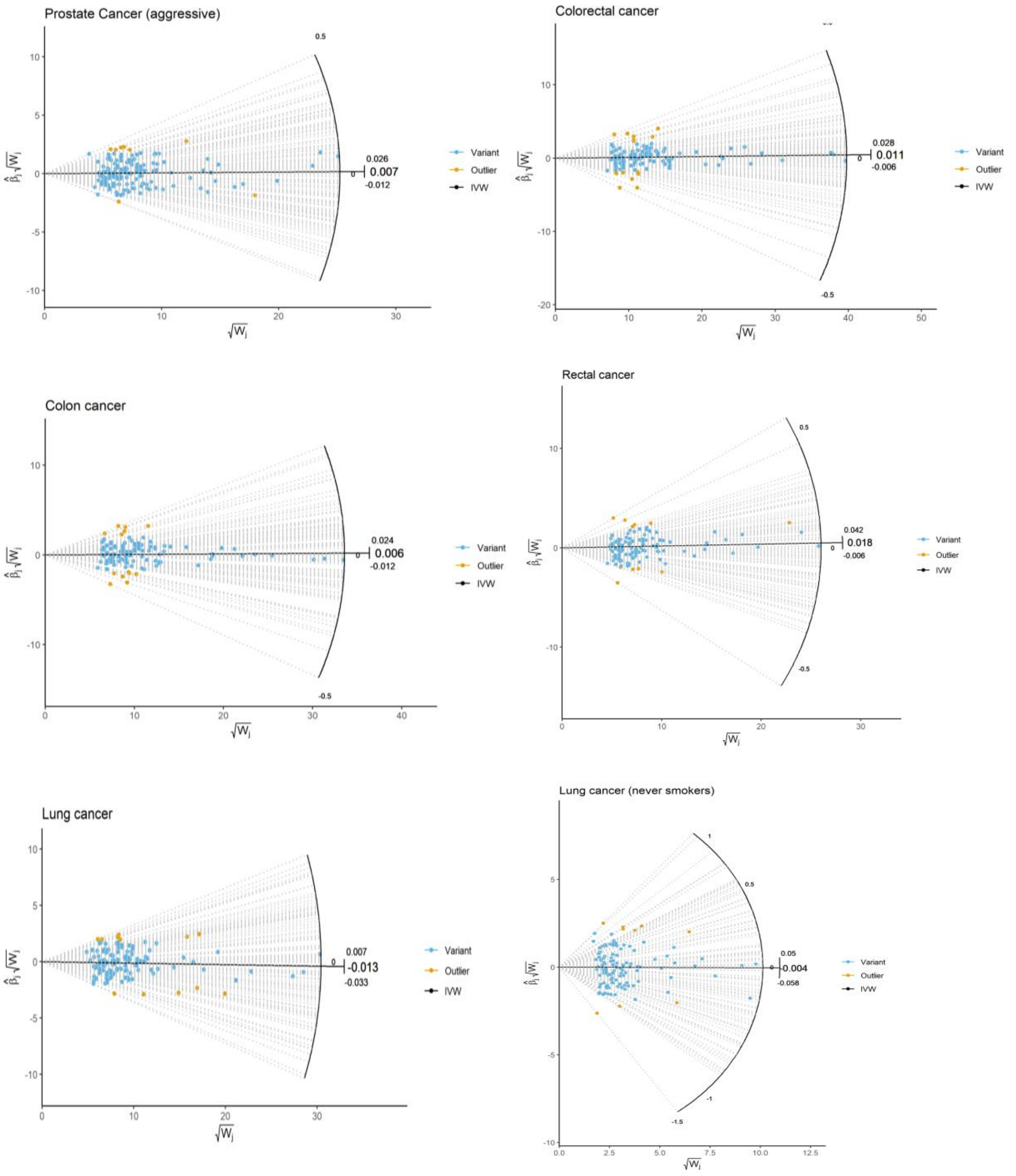


**Supplementary Figure 6: Radial plots of genetically predicted fitness with cancer outcomes (continued)**

Radial plots to visualize outlier SNPs in Mendelian randomization analysis. The radial curve shows the ratio estimate for each genetic variant and the IVW estimate. The lines join each data point back to the origin. Plots were produced using the RadialMR package[3].

Abbreviations: ER=estrogen receptor; IVW= inverse variance weighted; SNP=single nucleotide polymorphism.

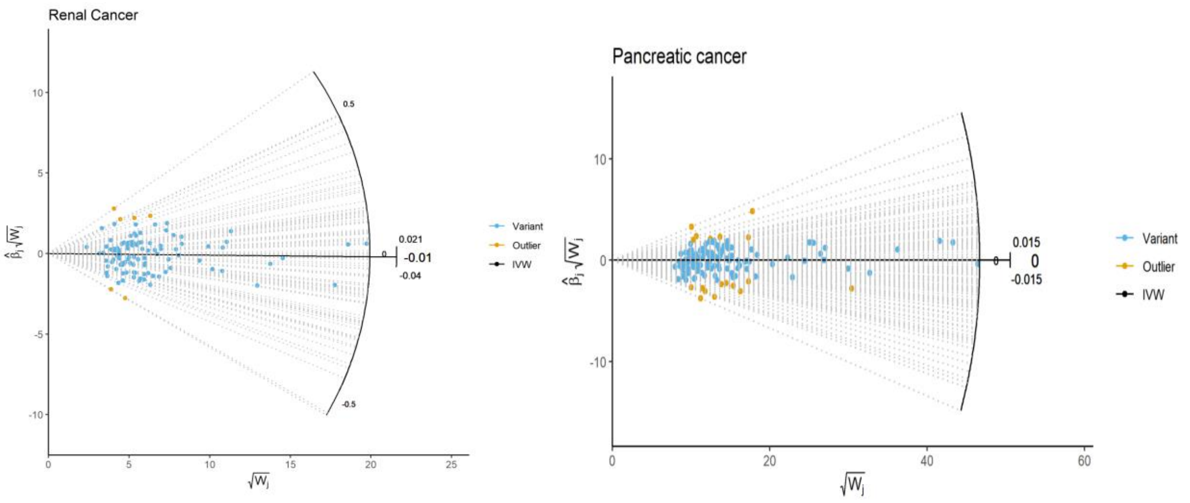


**Supplementary Figure 6: Radial plots of genetically predicted fitness with cancer outcomes (continued)**

Radial plots to visualize outlier SNPs in Mendelian randomization analysis. The radial curve shows the ratio estimate for each genetic variant and the IVW estimate. The lines join each data point back to the origin. Plots were produced using the RadialMR package. [3]

Abbreviations: IVW= inverse variance weighted; SNP=single nucleotide polymorphism.

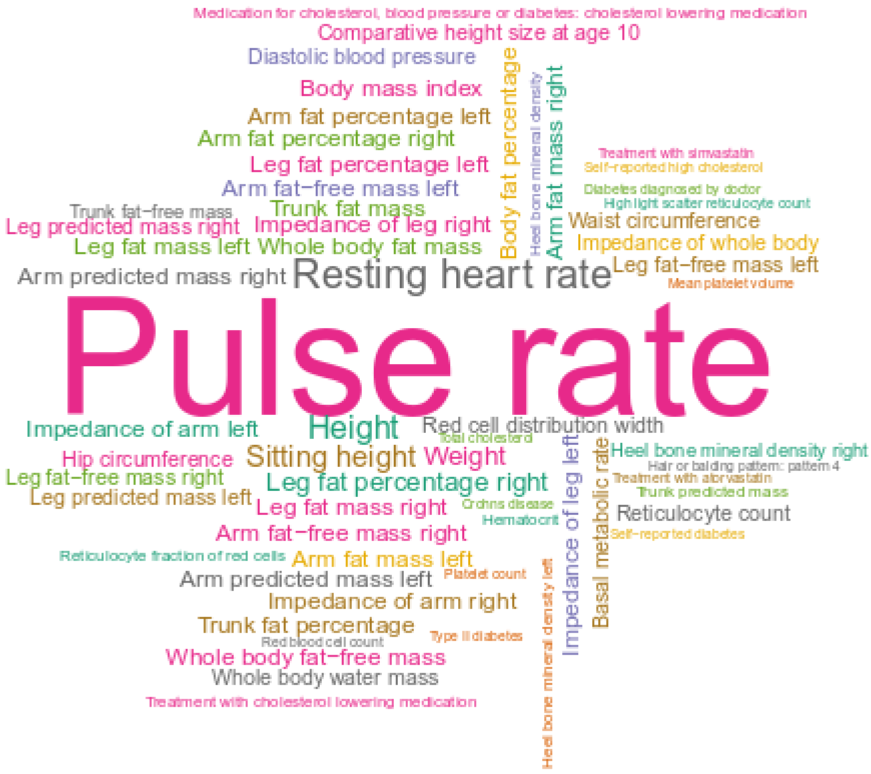


**Supplementary Figure 7: Traits associated with each SNP associated with fitness**

Traits were identified using the PhenoScanner resource. Larger words represent a greater frequency of the traits being associated with the SNPs (P-value threshold=5 x $10^{-8}$). This figure was created using the wordcloud package in R.

Abbreviations: SNP=single nucleotide polymorphism

**Supplementary Table 1: Genetic consortia case control numbers and further information**

| Site | Name | N cases | N controls | Reference |
|---|---|---|---|---|
| Breast | BCAC | 133,384 | 113,789 | Zhang, H. et al. Genome-wide association study identifies 32 novel breast cancer susceptibility loci from overall and subtype-specific analyses. Nat Genet 2020. doi: 10.1038/s41588-020-0609-2 |
| ER+ | BCAC | 69,501 | 105 974 | Michailidou, K. et al. Association analysis identifies 65 new breast cancer risk loci. Nature 2017. doi:10.1038/nature24285 |
| ER- | BCAC | 21,468 | 105 974 | |
| Colorectal | GECCO, CORECT, CCFR | 58,131 | 67,347 | Huyghe, J.R., et al. Discovery of common and rare genetic risk variants for colorectal cancer. Nat Genet 2019. doi: 10.1038/s41588-018-0286-6 |
| Male | | 31,288 | 34,527 | |
| Female | | 26,843 | 32,820 | |
| Colon | | 32,002 | 64,159 | Huyghe J.R., et al. Genetic architectures of proximal and distal colorectal cancer are partly distinct. Gut 2021. doi: 10.1136/gutjnl-2020-321534 |
| Rectal | | 16,212 | 64,159 | |
| Proximal colon | | 15,706 | 64,159 | |
| Distal colon | | 14,376 | 64,159 | |
| Endometrium | ECAC, E2C2, UK Biobank | 12,906 | 108,979 | O'Mara T et al. Identification of nine new susceptibility loci for endometrial cancer. Nat Comm 2018. doi: 10.1038/s41467-018-05427-7 |
| Lung | TRICL-ILCCO, LC3 | 29,266 | 56,450 | Mckay, J., et al. Large-scale association analysis identifies new lung cancer susceptibility loci and heterogeneity in genetic susceptibility across histological subtypes. Nat Genet 2017. doi: 10.1038/ng.3892 |
| Never-smokers | | 2,355 | 7,504 | |
| Ovary | OCAC | 22,406 | 40,941 | Phelan, C. M. et al. Identification of 12 new susceptibility loci for different histotypes of epithelial ovarian cancer. Nat Genet 2017. doi:10.1038/ng.3826; invasive cases. |
| Pancreatic | PanScan, PanC4 | 7,110 | 7,264 | Amundadottir L et al. Genome-wide association study identifies variants in the ABO locus associated with susceptibility to pancreatic cancer. Nat Genet 2009. doi: 10.1038/ng.429. Petersen GM et al. A genome-wide association study identifies pancreatic cancer susceptibility loci on chromosomes 13q22.1, 1q32.1 and 5p15.33. Nat Genet 2010. doi: 10.1038/ng.522 Childs EJ et al Common variation at 2p13.3, 3q29, 7p13 and 17q25.1 associated with susceptibility to pancreatic cancer. Nat Genet 2015. doi: 10.1038/ng.3341 |
| Prostate | PRACTICAL, GAME-ON/ ELLIPSE | 79,148 | 61,106 | Schumacher, F. R. et al. Association analyses of more than 140,000 men identify 63 new prostate cancer susceptibility loci. Nat Genet 2018. doi: 10.1038/s41588-018-0142-8 |
| Aggressive prostate* | | 15,167 | 58,308 | |
| Renal cell carcinoma | - | 10,784 | 20,406 | Scelo G et al. Genome-wide association study identifies multiple risk loci for renal cell carcinoma. Nature Comm 2017. doi: 10.1038/ncomms15724 |

*defined as: disease metastases at diagnosis, Gleason score 8+, prostate cancer death, or prostate-specific antigen (PSA) >100 ng/mL.

Abbreviations: BCAC=Breast Cancer Association Consortium; CCFR=Colon Cancer Family Registry; CORECT=Colorectal Cancer Transdisciplinary Study; E2C2= Epidemiology of Endometrial Cancer Consortium; ECAC=Endometrial Cancer Association Consortium; GECCO=Genetics and Epidemiology of Colorectal Cancer Consortium; LC3= Lung Cancer Cohort Consortium; OCAC=Ovarian Cancer Association Consortium; PRACTICAL= Prostate Cancer Association Group to Investigate Cancer Associated Alterations in the Genome, TRICL-ILCCO = Transdisciplinary Research of Cancer in Lung of the International Lung Cancer Consortium ;

### Supplementary Table 2: Participant characteristics by age-adjusted and sex-specific cardiorespiratory fitness ($VO_2$max per kg fat-free mass) tertiles

| | Women | | | Men | | |
|---|---|---|---|---|---|---|
| | Lower fitness | Mid fitness | Higher fitness | Lower fitness | Mid fitness | Higher fitness |
| N | 12791 | 12790 | 12786 | 11404 | 11402 | 11399 |
| Age (y) | 57 ± 8 | 57 ± 8 | 57 ± 8 | 58 ± 8 | 58 ± 8 | 58 ± 8 |
| Height (m) | 1.63 ± 0.06 | 1.63 ± 0.06 | 1.63 ± 0.06 | 1.77 ± 0.07 | 1.76 ± 0.07 | 1.75 ± 0.07 |
| Total-body mass (kg) | 73.3 ± 14.8 | 70.7 ± 12.9 | 67.7 ± 11.4 | 90.8 ± 14.8 | 85.2 ± 12.5 | 80.2 ± 11.3 |
| Fat-free mass (kg) | 45.3 ± 5.3 | 44.3 ± 4.7 | 43.4 ± 4.3 | 66.3 ± 7.8 | 63.5 ± 7.1 | 60.9 ± 6.7 |
| BMI ($kg \cdot m^{-2}$) | 27.7 ± 5.4 | 26.6 ± 4.6 | 25.4 ± 4.1 | 29.1 ± 4.4 | 27.5 ± 3.6 | 26.1 ± 3.3 |
| $VO_2max_{tbm}$($ml \cdot min^{-1} \cdot kg^{-1}$) | 20.5 ± 3.9 | 25.2 ± 3.2 | 30.5 ± 5.3 | 26.5 ± 3.5 | 31.8 ± 3.0 | 37.8 ± 4.7 |
| $VO_2max_{ffm}$ ($ml \cdot min^{-1} \cdot kg^{-1}$) | 32.5 ± 5.0 | 39.5 ± 2.3 | 46.8 ± 5.5 | 35.9 ± 3.5 | 42.3 ± 2.1 | 49.4 ± 4.3 |
| Red meat consumption | 0.8 ± 0.5 | 0.8 ± 0.5 | 0.7 ± 0.5 | 1.1 ± 0.6 | 1.0 ± 0.6 | 1.0 ± 0.6 |
| Fish consumption | | | | | | |
| Never | 10.3% (1319) | 9.3% (1192) | 8.4% (1073) | 11.5% (1313) | 11.0% (1249) | 9.2% (1052) |
| At most 1 per week | 32.4% (4144) | 32.5% (4163) | 31.6% (4046) | 35.9% (4096) | 33.9% (3865) | 32.8% (3742) |
| 2 or more per week | 56.5% (7230) | 57.6% (7370) | 59.6% (7621) | 51.5% (5875) | 54.6% (6221) | 57.4% (6546) |
| Missing | 0.8% (98) | 0.5% (65) | 0.4% (46) | 1.1% (120) | 0.6% (67) | 0.5% (59) |
| Fruit & vegetable consumption | | | | | | |
| Never | 16.6% (2125) | 15.6% (1998) | 13.5% (1722) | 24.7% (2815) | 22.8% (2599) | 20.5% (2332) |
| At most 1 per week | 29.1% (3716) | 28.1% (3588) | 26.6% (3401) | 33.3% (3800) | 32.3% (3687) | 31.7% (3618) |
| 2 or more per week | 54.0% (6901) | 56.1% (7175) | 59.8% (7644) | 41.5% (4736) | 44.6% (5086) | 47.6% (5422) |
| Missing | 0.4% (49) | 0.2% (29) | 0.1% (19) | 0.5% (53) | 0.3% (30) | 0.2% (27) |
| Salt addition to meals | | | | | | |
| Never/rarely | 59.3% (7585) | 57.8% (7392) | 58.2% (7443) | 54.5% (6210) | 56.0% (6386) | 58.6% (6685) |
| Sometimes | 26.5% (3390) | 27.4% (3508) | 27.7% (3544) | 28.3% (3232) | 28.0% (3196) | 26.6% (3031) |
| Usually/always | 13.9% (1773) | 14.6% (1869) | 13.9% (1783) | 16.9% (1923) | 15.8% (1800) | 14.6% (1665) |
| Missing | 0.3% (43) | 0.2% (21) | 0.1% (16) | 0.3% (39) | 0.2% (20) | 0.2% (18) |
| Alcohol consumption | | | | | | |
| Never or previous | 11.0% (1410) | 8.2% (1050) | 6.6% (846) | 6.9% (787) | 6.0% (686) | 4.9% (559) |
| At most 2 per week | 57.1% (7302) | 53.0% (6777) | 49.4% (6312) | 45.8% (5221) | 42.1% (4799) | 39.0% (4451) |
| 3 or more per week | 31.5% (4032) | 38.6% (4935) | 43.9% (5607) | 46.9% (5349) | 51.7% (5895) | 55.9% (6368) |
| Missing | 0.4% (47) | 0.2% (28) | 0.2% (21) | 0.4% (47) | 0.2% (22) | 0.2% (21) |
| Smoking status | | | | | | |
| Never | 65.0% (8314) | 60.7% (7760) | 59.3% (7577) | 49.5% (5642) | 51.8% (5907) | 54.0% (6157) |
| Previous | 27.6% (3530) | 31.8% (4073) | 33.3% (4261) | 39.0% (4447) | 37.8% (4306) | 36.4% (4146) |
| Current | 6.8% (870) | 7.0% (897) | 7.1% (902) | 10.8% (1231) | 9.9% (1130) | 9.2% (1045) |
| Missing | 0.6% (77) | 0.5% (60) | 0.4% (46) | 0.7% (84) | 0.5% (59) | 0.4% (51) |
| Townsend deprivation index | -1.1 ± 3.0 | -1.3 ± 2.9 | -1.4 ± 2.8 | -1.0 ± 3.1 | -1.3 ± 2.9 | -1.5 ± 2.9 |
| Education | | | | | | |
| No qualification | 13.8% (1763) | 10.5% (1346) | 9.4% (1204) | 14.0% (1601) | 12.4% (1414) | 9.8% (1119) |
| Any other qualification | 52.3% (6689) | 51.0% (6520) | 48.3% (6178) | 50.9% (5802) | 48.5% (5526) | 44.2% (5037) |
| Degree level or above | 32.5% (4156) | 37.6% (4810) | 41.6% (5323) | 33.6% (3834) | 38.2% (4358) | 45.2% (5153) |
| Missing | 1.4% (183) | 0.9% (114) | 0.6% (81) | 1.5% (167) | 0.9% (104) | 0.8% (90) |
| Employment | | | | | | |
| Unemployed | 9.9% (1268) | 8.2% (1054) | 7.9% (1015) | 7.9% (900) | 5.7% (651) | 4.9% (560) |
| Employed | 53.1% (6786) | 56.0% (7163) | 57.1% (7305) | 56.9% (6492) | 60.2% (6865) | 61.2% (6975) |
| Retired | 36.2% (4636) | 35.3% (4512) | 34.6% (4428) | 34.4% (3924) | 33.5% (3814) | 33.4% (3808) |
| Missing | 0.8% (101) | 0.5% (61) | 0.3% (38) | 0.8% (88) | 0.6% (72) | 0.5% (56) |
| Race | | | | | | |
| Asian or Asian British | 3.2% (406) | 2.3% (291) | 1.8% (232) | 3.3% (371) | 3.4% (389) | 2.8% (322) |
| Black or Black British | 4.7% (602) | 2.4% (304) | 1.3% (162) | 3.6% (411) | 2.0% (232) | 1.3% (147) |
| Mixed | 0.9% (120) | 1.0% (131) | 0.8% (104) | 0.7% (77) | 0.7% (79) | 0.7% (77) |
| Other | 2.3% (294) | 1.7% (213) | 1.6% (210) | 1.7% (193) | 1.5% (175) | 1.5% (166) |
| White | 88.1% (11271) | 92.2% (11791) | 94.0% (12019) | 90.0% (10262) | 91.7% (10458) | 93.2% (10619) |
| Missing | 0.8% (98) | 0.5% (60) | 0.5% (59) | 0.8% (90) | 0.6% (69) | 0.6% (68) |
| Hypertension | | | | | | |
| Not hypertensive | 43.9% (5614) | 56.1% (7173) | 63.8% (8162) | 28.9% (3294) | 41.6% (4744) | 52.4% (5972) |
| Hypertensive | 56.1% (7177) | 43.9% (5617) | 36.2% (4624) | 71.1% (8110) | 58.4% (6658) | 47.6% (5427) |
| Diabetes | | | | | | |
| Not diabetic | 94.1% (12041) | 97.0% (12410) | 98.1% (12538) | 88.5% (10091) | 94.2% (10745) | 96.6% (11012) |
| Diabetic | 5.5% (698) | 2.8% (356) | 1.8% (234) | 11.1% (1269) | 5.6% (638) | 3.3% (371) |
| Missing | 0.4% (52) | 0.2% (24) | 0.1% (14) | 0.4% (44) | 0.2% (19) | 0.1% (16) |
| Colorectal cancer | 0.9% (118) | 0.9% (121) | 0.8% (104) | 1.5% (167) | 1.4% (165) | 1.2% (136) |
| Colon | 0.7% (90) | 0.8% (98) | 0.6% (82) | 1.0% (116) | 1.1% (121) | 0.8% (91) |
| Rectal | 0.3% (42) | 0.3% (37) | 0.3% (42) | 0.7% (84) | 0.6% (72) | 0.6% (71) |
| Lung cancer | 0.6% (77) | 0.6% (77) | 0.6% (76) | 0.9% (101) | 0.7% (82) | 0.6% (67) |
| Breast cancer | 3.2% (404) | 2.7% (344) | 2.7% (345) | | | |
| Endometrial cancer | 0.6% (73) | 0.5% (66) | 0.4% (45) | | | |
| Ovarian cancer | 0.3% (38) | 0.4% (52) | 0.4% (46) | | | |
| Prostate cancer | | | | 4.3% (489) | 5.0% (574) | 4.6% (523) |

**Supplementary Table 3: Associations of cardiorespiratory respiratory fitness and cancer risk in minimally adjusted models**

| Cancer site | HR per 3.5 ml $O_2 \cdot min^{-1} \cdot kg^{-1}$ total-body mass | HR per 5.0 ml $O_2 \cdot min^{-1} \cdot kg^{-1}$ fat-free mass |
|---|---|---|
| Colorectal | 0.93 (0.89, 0.98) | 0.96 (0.91, 1.01) |
| Colon | 0.94 (0.89, 0.99) | 0.97 (0.91, 1.03) |
| Rectal | 0.95 (0.88, 1.01) | 0.96 (0.89, 1.04) |
| Lung | 0.97 (0.92, 1.03) | 0.97 (0.90, 1.04) |
| Nonsmokers | 0.96 (0.84, 1.09) | 0.97 (0.83, 1.12) |
| Breast | 0.97 (0.93, 1.00) | 0.98 (0.94, 1.02) |
| Endometrial | 0.81 (0.73, 0.89) | 0.92 (0.83, 1.02) |
| Ovarian | 1.06 (0.96, 1.18) | 1.08 (0.96, 1.22) |
| Prostate | 1.03 (1.00, 1.06) | 1.03 (1.00, 1.07) |

HR and 95% CI for fitness and cancer diagnosis. Analyses were adjusted for age and sex

Abbreviations: CI=confidence interval; HR=hazard ratio.

**Supplementary Table 4: Associations per genetically predicted 5.0 ml $O_2 \cdot min^{-1} \cdot kg^{-1}$ increment in fat-free mass and colorectal cancer risk by sex**

| | Males | | Females | |
|---|---|---|---|---|
| | OR (95% CI) | P-value | OR (95% CI) | P-value |
| Inverse variance weighted | 1.08 (0.98, 1.2) | 0.13 | 1.02 (0.92, 1.13) | 0.68 |
| Weighted median | 1.06 (0.92, 1.22) | 0.41 | 1.01 (0.88, 1.16) | 0.89 |
| MR-PRESSO | 1.09 (0.99, 1.2) | 0.08 | 1.01 (0.92, 1.12) | 0.79 |
| Contamination mixture | 1.14 (0.98, 1.26) | 0.15 | 1.00 (0.91, 1.11) | 0.92 |

Abbreviations: CI=confidence interval; MR=Mendelian randomization; OR=odds ratio; PRESSO=pleiotropy residual sum and outlier.

**Supplementary Table 5: Associations per genetically predicted 5.0 ml $O_2 \cdot min^{-1} \cdot kg^{-1}$ increment in fat-free mass and colorectal cancer risk by site (proximal/distal)**

| | Proximal | | Distal | |
|---|---|---|---|---|
| | OR (95% CI) | P-value | OR (95% CI) | P-value |
| Inverse variance weighted | 1.00 (0.90, 1.13) | 0.95 | 1.04 (0.93, 1.16) | 0.51 |
| Weighted median | 0.95 (0.81, 1.11) | 0.52 | 0.99 (0.85, 1.16) | 0.92 |
| MR-PRESSO | 1.00 (0.90, 1.13) | 0.95 | 1.03 (0.92, 1.15) | 0.63 |
| Contamination mixture | 0.94 (0.85, 1.15) | 0.48 | 1.05 (0.95, 1.16) | 0.78 |

Abbreviations: CI=confidence interval; MR=Mendelian randomization; OR=odds ratio; PRESSO=pleiotropy residual sum and outlier.

**Supplementary Table 6**

Mendelian randomization sensitivity analyses per genetically predicted 5.0 ml $O_2 \cdot min^{-1} \cdot kg^{-1}$ fat-free mass

| | Weighted median | | MR-PRESSO | | Contamination mixture | |
|---|---|---|---|---|---|---|
| Cancer site | OR (95% CI) | P-value | OR (95% CI) | P-value | OR (95%CI) | P-value |
| Breast | 0.99 (0.92, 1.06) | 0.73 | **0.94 (0.89, 0.99)** | **0.03** | 0.96 (0.91, 1.01) | 0.41 |
| ER - | 0.92 (0.81, 1.05) | 0.23 | **0.88 (0.80, 0.97)** | **0.01** | **0.86 (0.78, 0.95)** | **0.03** |
| ER + | 0.96 (0.88, 1.06) | 0.44 | **0.93 (0.87, 0.99)** | **0.03** | 0.99 (0.94, 1.04) | 1.00 |
| Lung | **0.85 (0.74, 0.98)** | **0.03** | 0.94 (0.85, 1.03) | 0.18 | 0.88 (0.72, 0.97) | 0.07 |
| Never smokers | 1.05 (0.69, 1.60) | 0.82 | 0.98 (0.75, 1.28) | 0.88 | 0.71 (0.47, 1.23) | 0.25 |
| Endometrial | 1.08 (0.91, 1.27) | 0.37 | 0.95 (0.84, 1.07) | 0.38 | 1.07 (0.83, 1.31) | 0.39 |
| Renal | 0.97 (0.79, 1.19) | 0.77 | 0.95 (0.83, 1.10) | 0.50 | 0.92 (0.75, 1.18) | 0.69 |
| Ovarian | 0.97 (0.84, 1.13) | 0.71 | 0.91 (0.83, 1.00) | 0.05 | 0.98 (0.85, 1.03) | 0.56 |
| Prostate | 1.00 (0.91, 1.10) | 0.98 | 1.00 (0.93, 1.06) | 0.89 | 1.06 (1.01, 1.11) | 0.16 |
| Aggressive | 1.08 (0.92, 1.27) | 0.32 | 1.03 (0.94, 1.14) | 0.49 | 0.86 (0.74, 1.22) | 0.28 |
| Pancreatic | 1.01 (0.93, 1.11) | 0.75 | 1.00 (0.94, 1.07) | 0.96 | **1.09 (1.03, 1.14)** | **0.03** |
| Colorectal | 1.06 (0.96, 1.17) | 0.24 | 1.05 (0.98, 1.12) | 0.18 | 1.05 (1.00, 1.11) | 0.17 |
| Colon | 0.97 (0.86, 1.10) | 0.65 | 1.03 (0.94, 1.12) | 0.58 | 1.05 (0.95, 1.1) | 0.63 |
| Rectal | 1.11 (0.94, 1.31) | 0.20 | 1.10 (0.97, 1.24) | 0.12 | 1.16 (1.00, 1.35) | 0.11 |

Risk estimates $p<0.05$ are in bold.

Abbreviations: CI=confidence interval; MR=Mendelian randomization; OR=odds ratio; PRESSO=pleiotropy residual sum and outlier.

**References**


1. UK Biobank cardio assessment manual: Version 1.0. Published online April 5, 2011. http://biobank.ctsu.ox.ac.uk/crystal/docs/Cardio.pdf

2. Gonzales TI, Westgate K, Strain T, et al. Cardiorespiratory fitness assessment using risk-stratified exercise testing and dose-response relationships with disease outcomes. *Sci Rep*. 2021;11(1):15315. doi:10.1038/s41598-021-94768-3

3. Bowden J, Spiller W, Del Greco M F, et al. Improving the visualization, interpretation and analysis of two-sample summary data Mendelian randomization via the Radial plot and Radial regression. *Int J Epidemiol*. 2018;47(4):1264-1278. doi:10.1093/ije/dyy101
