## Appendix 1 for "Observational and genetic associations between cardiorespiratory fitness and cancer: a UK Biobank and international consortia study"

**Appendix 1: PIs from the PRACTICAL (http://practical.icr.ac.uk/), CRUK, BPC3, CAPS, PEGASUS consortia**

Rosalind A. Eeles[1,2], Christopher A. Haiman[3], Zsofia Kote-Jarai[1], Fredrick R. Schumacher[4,5], Sara Benlloch[6,1], Ali Amin Al Olama[6,7], Kenneth R. Muir[8], Sonja I. Berndt[9], David V. Conti[3], Fredrik Wiklund[10], Stephen Chanock[9], Ying Wang[11], Catherine M. Tangen[12], Jyotsna Batra[13,14], Judith A. Clements[13,14], APCB BioResource (Australian Prostate Cancer BioResource)[15,14], Henrik Grönberg[10], Nora Pashayan[16,17], Johanna Schleutker[18,19], Demetrius Albanes[9], Stephanie J. Weinstein[9], Alicja Wolk[20], Catharine M. L. West[21], Lorelei A. Mucci[22], Géraldine Cancel-Tassin[23,24], Stella Koutros[9], Karina Dalsgaard Sørensen[25,26], Eli Marie Grindedal[27], David E. Neal[28,29,30], Freddie C. Hamdy[31,32], Jenny L. Donovan[33], Ruth C. Travis[34], Robert J. Hamilton[35,36], Sue Ann Ingles[37], Barry S. Rosenstein[38], Yong-Jie Lu[39], Graham G. Giles[40,41,42], Robert J. MacInnis[40,41], Adam S. Kibel[43], Ana Vega[44,45,46], Manolis Kogevinas[47,48,49,50], Kathryn L. Penney[51], Jong Y. Park[52], Janet L. Stanford[53,54], Cezary Cybulski[55], Børge G. Nordestgaard[56,57], Sune F. Nielsen[56,57], Hermann Brenner[58,59,60], Christiane Maier[61], Jeri Kim[62], Esther M. John[63], Manuel R. Teixeira[64,65,66], Susan L. Neuhausen[67], Kim De Ruyck[68], Azad Razack[69], Lisa F. Newcomb[53,70], Davor Lessel[71], Radka Kaneva[72], Nawaid Usmani[73,74], Frank Claessens[75], Paul A. Townsend[76,77], Jose Esteban Castelao[78], Monique J. Roobol[79], Florence Menegaux[80], Kay-Tee Khaw[81], Lisa Cannon-Albright[82,83], Hardev Pandha[77], Stephen N. Thibodeau[84], David J. Hunter[85], Peter Kraft[86], William J. Blot[87,88], Elio Riboli[89]

[1]The Institute of Cancer Research, London, SM2 5NG, UK
[2]Royal Marsden NHS Foundation Trust, London, SW3 6JJ, UK
[3]Center for Genetic Epidemiology, Department of Preventive Medicine, Keck School of Medicine, University of Southern California/Norris Comprehensive Cancer Center, Los Angeles, CA 90015, USA
[4]Department of Population and Quantitative Health Sciences, Case Western Reserve University, Cleveland, OH 44106-7219, USA
[5]Seidman Cancer Center, University Hospitals, Cleveland, OH 44106, USA.
[6]Centre for Cancer Genetic Epidemiology, Department of Public Health and Primary Care, University of Cambridge, Strangeways Research Laboratory, Cambridge CB1 8RN, UK
[7]University of Cambridge, Department of Clinical Neurosciences, Stroke Research Group, R3, Box 83, Cambridge Biomedical Campus, Cambridge CB2 0QQ, UK
[8]Division of Population Health, Health Services Research and Primary Care, University of Manchester, Oxford Road, Manchester, M13 9PL, UK
[9]Division of Cancer Epidemiology and Genetics, National Cancer Institute, NIH, Bethesda, Maryland, 20892, USA
[10]Department of Medical Epidemiology and Biostatistics, Karolinska Institute, SE-171 77 Stockholm, Sweden

[11]Department of Population Science, American Cancer Society, 250 Williams Street, Atlanta, GA 30303, USA
[12]SWOG Statistical Center, Fred Hutchinson Cancer Research Center, Seattle, WA 98109, USA
[13]Australian Prostate Cancer Research Centre-Qld, Institute of Health and Biomedical Innovation and School of Biomedical Sciences, Queensland University of Technology, Brisbane QLD 4059, Australia
[14]Translational Research Institute, Brisbane, Queensland 4102, Australia
[15]Australian Prostate Cancer Research Centre-Qld, Queensland University of Technology, Brisbane; Prostate Cancer Research Program, Monash University, Melbourne; Dame Roma Mitchell Cancer Centre, University of Adelaide, Adelaide; Chris O'Brien Lifehouse and The Kinghorn Cancer Centre, Sydney, Australia
[16]Department of Applied Health Research, University College London, London, WC1E 7HB, UK
[17]Centre for Cancer Genetic Epidemiology, Department of Oncology, University of Cambridge, Strangeways Laboratory, Worts Causeway, Cambridge, CB1 8RN, UK
[18]Institute of Biomedicine, University of Turku, Finland
[19]Department of Medical Genetics, Genomics, Laboratory Division, Turku University Hospital, PO Box 52, 20521 Turku, Finland
[20]Department of Surgical Sciences, Uppsala University, 75185 Uppsala, Sweden
[21]Division of Cancer Sciences, University of Manchester, Manchester Academic Health Science Centre, Radiotherapy Related Research, The Christie Hospital NHS Foundation Trust, Manchester, M13 9PL UK
[22]Department of Epidemiology, Harvard T. H. Chan School of Public Health, Boston, MA 02115, USA
[23]CeRePP, Tenon Hospital, F-75020 Paris, France.
[24]Sorbonne Universite, GRC n°5 , AP-HP, Tenon Hospital, 4 rue de la Chine, F-75020 Paris, France
[25]Department of Molecular Medicine, Aarhus University Hospital, Palle Juul-Jensen Boulevard 99, 8200 Aarhus N, Denmark
[26]Department of Clinical Medicine, Aarhus University, DK-8200 Aarhus N
[27]Department of Medical Genetics, Oslo University Hospital, 0424 Oslo, Norway
[28]Nuffield Department of Surgical Sciences, University of Oxford, Room 6603, Level 6, John Radcliffe Hospital, Headley Way, Headington, Oxford, OX3 9DU, UK
[29]University of Cambridge, Department of Oncology, Box 279, Addenbrooke's Hospital, Hills Road, Cambridge CB2 0QQ, UK
[30]Cancer Research UK, Cambridge Research Institute, Li Ka Shing Centre, Cambridge, CB2 0RE, UK
[31]Nuffield Department of Surgical Sciences, University of Oxford, Oxford, OX1 2JD, UK
[32]Faculty of Medical Science, University of Oxford, John Radcliffe Hospital, Oxford, UK
[33]Population Health Sciences, Bristol Medical School, University of Bristol, BS8 2PS, UK
[34]Cancer Epidemiology Unit, Nuffield Department of Population Health, University of Oxford, Oxford, OX3 7LF, UK
[35]Dept. of Surgical Oncology, Princess Margaret Cancer Centre, Toronto ON M5G 2M9,

Canada
[36]Dept. of Surgery (Urology), University of Toronto, Canada
[37]Department of Preventive Medicine, Keck School of Medicine, University of Southern California/Norris Comprehensive Cancer Center, Los Angeles, CA 90015, USA
[38]Department of Radiation Oncology and Department of Genetics and Genomic Sciences, Box 1236, Icahn School of Medicine at Mount Sinai, One Gustave L. Levy Place, New York, NY 10029, USA
[39]Centre for Cancer Biomarker and Biotherapeutics, Barts Cancer Institute, Queen Mary University of London, John Vane Science Centre, Charterhouse Square, London, EC1M 6BQ, UK
[40]Cancer Epidemiology Division, Cancer Council Victoria, 615 St Kilda Road, Melbourne, VIC 3004, Australia
[41]Centre for Epidemiology and Biostatistics, Melbourne School of Population and Global Health, The University of Melbourne, Grattan Street, Parkville, VIC 3010, Australia
[42]Precision Medicine, School of Clinical Sciences at Monash Health, Monash University, Clayton, Victoria 3168, Australia
[43]Division of Urologic Surgery, Brigham and Womens Hospital, 75 Francis Street, Boston, MA 02115, USA
[44]Fundación Pública Galega Medicina Xenómica, Santiago de Compostela, 15706, Spain.
[45]Instituto de Investigación Sanitaria de Santiago de Compostela, Santiago de Compostela, 15706, Spain.
[46]Centro de Investigación en Red de Enfermedades Raras (CIBERER), Spain
[47]ISGlobal, Barcelona, Spain
[48]IMIM (Hospital del Mar Medical Research Institute), Barcelona, Spain
[49]Universitat Pompeu Fabra (UPF), Barcelona, Spain
[50]CIBER Epidemiología y Salud Pública (CIBERESP), 28029 Madrid, Spain
[51]Channing Division of Network Medicine, Department of Medicine, Brigham and Women's Hospital/Harvard Medical School, Boston, MA 02115, USA
[52]Department of Cancer Epidemiology, Moffitt Cancer Center, 12902 Magnolia Drive, Tampa, FL 33612, USA
[53]Division of Public Health Sciences, Fred Hutchinson Cancer Research Center, Seattle, Washington, 98109-1024, USA
[54]Department of Epidemiology, School of Public Health, University of Washington, Seattle, Washington 98195, USA
[55]International Hereditary Cancer Center, Department of Genetics and Pathology, Pomeranian Medical University, 70-115 Szczecin, Poland
[56]Faculty of Health and Medical Sciences, University of Copenhagen, 2200 Copenhagen, Denmark
[57]Department of Clinical Biochemistry, Herlev and Gentofte Hospital, Copenhagen University Hospital, Herlev, 2200 Copenhagen, Denmark
[58]Division of Clinical Epidemiology and Aging Research, German Cancer Research Center (DKFZ), D-69120, Heidelberg, Germany
[59]German Cancer Consortium (DKTK), German Cancer Research Center (DKFZ), D-69120 Heidelberg, Germany

[60]Division of Preventive Oncology, German Cancer Research Center (DKFZ) and National Center for Tumor Diseases (NCT), Im Neuenheimer Feld 460, 69120 Heidelberg, Germany
[61]Humangenetik Tuebingen, Paul-Ehrlich-Str 23, D-72076 Tuebingen, Germany
[62]The University of Texas M. D. Anderson Cancer Center, Department of Genitourinary Medical Oncology, 1515 Holcombe Blvd., Houston, TX 77030, USA
[63]Departments of Epidemiology & Population Health and of Medicine, Division of Oncology, Stanford Cancer Institute, Stanford University School of Medicine, Stanford, CA 94304 USA
[64]Department of Laboratory Genetics, Portuguese Oncology Institute of Porto (IPO Porto) / Porto Comprehensive Cancer Center, Porto, Portugal
[65]Cancer Genetics Group, IPO Porto Research Center (CI-IPOP) / RISE@CI-IPOP (Health Research Network), Portuguese Oncology Institute of Porto (IPO Porto) / Porto Comprehensive Cancer Center, Porto, Portugal
[66]School of Medicine and Biomedical Sciences (ICBAS), University of Porto, Porto, Portugal
[67]Department of Population Sciences, Beckman Research Institute of the City of Hope, 1500 East Duarte Road, Duarte, CA 91010
[68]Ghent University, Faculty of Medicine and Health Sciences, Basic Medical Sciences, Proeftuinstraat 86, B-9000 Gent
[69]Department of Surgery, Faculty of Medicine, University of Malaya, 50603 Kuala Lumpur, Malaysia
[70]Department of Urology, University of Washington, 1959 NE Pacific Street, Box 356510, Seattle, WA 98195, USA
[71]Institute of Human Genetics, University Medical Center Hamburg-Eppendorf, D-20246 Hamburg, Germany
[72]Molecular Medicine Center, Department of Medical Chemistry and Biochemistry, Medical University of Sofia, Sofia, 2 Zdrave Str., 1431 Sofia, Bulgaria
[73]Department of Oncology, Cross Cancer Institute, University of Alberta, 11560 University Avenue, Edmonton, Alberta, Canada T6G 1Z2
[74]Division of Radiation Oncology, Cross Cancer Institute, 11560 University Avenue, Edmonton, Alberta, Canada T6G 1Z2
[75]Molecular Endocrinology Laboratory, Department of Cellular and Molecular Medicine, KU Leuven, BE-3000, Belgium
[76]Division of Cancer Sciences, Manchester Cancer Research Centre, Faculty of Biology, Medicine and Health, Manchester Academic Health Science Centre, NIHR Manchester Biomedical Research Centre, Health Innovation Manchester, Univeristy of Manchester, M13 9WL
[77]The University of Surrey, Guildford, Surrey, GU2 7XH, UK
[78]Genetic Oncology Unit, CHUVI Hospital, Complexo Hospitalario Universitario de Vigo, Instituto de Investigación Biomédica Galicia Sur (IISGS), 36204, Vigo (Pontevedra), Spain
[79]Department of Urology, Erasmus University Medical Center, Cancer Institute, 3015 GD Rotterdam, The Netherlands
[80]"Exposome and Heredity", CESP (UMR 1018), Faculté de Médecine, Université Paris-Saclay, Inserm, Gustave Roussy, Villejuif
[81]Clinical Gerontology Unit, University of Cambridge, Cambridge, CB2 2QQ, UK
[82]Division of Epidemiology, Department of Internal Medicine, University of Utah School of

Medicine, Salt Lake City, Utah 84132, USA
[83]George E. Wahlen Department of Veterans Affairs Medical Center, Salt Lake City, Utah 84148, USA
[84]Department of Laboratory Medicine and Pathology, Mayo Clinic, Rochester, MN 55905, USA
[85]Nuffield Department of Population Health, University of Oxford, United Kingdom
[86]Program in Genetic Epidemiology and Statistical Genetics, Department of Epidemiology, Harvard School of Public Health, Boston, MA, USA
[87]Division of Epidemiology, Department of Medicine, Vanderbilt University Medical Center, 2525 West End Avenue, Suite 800, Nashville, TN 37232 USA.
[88]International Epidemiology Institute, Rockville, MD 20850, USA
[89]Department of Epidemiology and Biostatistics, School of Public Health, Imperial College London, SW7 2AZ, UK
